# Association of Obesity with COVID-19 Severity and Mortality: A Systemic Review and Meta-Regression

**DOI:** 10.1101/2021.05.08.21256845

**Authors:** Romil Singh, Sawai Singh Rathore, Hira Khan, Smruti Karale, Abhishek Bhurwal, Aysun Tekin, Nirpeksh Jain, Ishita Mehra, Sohini Anand, Sanjana Reddy, Guneet Singh Sidhu, Anastasios Panagopoulos, Vishwanath Pattan, Rahul Kashyap, Vikas Bansal

**Author notes:** Corresponding Author: Vishwanath Pattan, MD Division of Endocrinology Wyoming Medical Center 419 S Washington St Suite 201 Casper, WY-82601. Conflict of Interest: None of the co-authors have anything to declare. Financial Support: There is no financial disclosure related to this study. **Author Contribution**: *Authors 1(RS) and 2(SSR) contributed equally in defining the study outline and manuscript writing. Data review and collection were done by AT, GSS, HK, NJ, RS, SK, AP, and SSR; statistical analysis was done by AB, SK, and VB; risk of bias was done by AT, SA and SSR. Study design, Distribution of Articles for critical review done by IM, VP, RK, and VB. Final approval received by all authors. RS, SSR, VB and VP is the guarantor of the paper, taking responsibility for the integrity of the work as a whole, from inception to published article. All authors contributed to the article and approved the submitted version.

## Abstract

**Objective:** To estimate the association of obesity with severity (defined as use of invasive mechanical ventilation or intensive care unit admission) and all-cause mortality in coronavirus disease 2019 (COVID-19) patients.

**Patients and Methods:** A systematic search was conducted from inception of COVID-19 pandemic through January 31st, 2021 for full-length articles focusing on the association of increased BMI/ Obesity and outcome in COVID-19 patients with help of various databases including Medline (PubMed), Embase, Science Web, and Cochrane Central Controlled Trials Registry. Preprint servers such as BioRxiv, MedRxiv, ChemRxiv, and SSRN were also scanned. Preferred Reporting Items for Systematic Reviews and Meta-Analysis (PRISMA) guidelines were used for study selection and data extraction. The severity in hospitalized COVID-19 patients, such as requirement of invasive mechanical ventilation and intensive care unit admission with high BMI/ Obesity was the chief outcome. While all-cause mortality in COVID-19 hospitalized patients with high BMI/ Obesity was the secondary outcome.

**Results:** A total of 576,784 patients from 100 studies were included in this meta-analysis. Being obese was associated with increased risk of severe disease (RR=1.46, 95% CI 1.34-1.60, p<0.001, I^2^ = 92 %). Similarly, high mortality was observed in obese patients with COVID-19 disease (RR=1.12, 95% CI 1.06-1.19, p<0.001, I^2^ = 88%). In a multivariate meta-regression on severity outcome, the covariate of female gender, pulmonary disease, diabetes, older age, cardiovascular diseases, and hypertension was found to be significant and explained R^2^= 50% of the between-study heterogeneity for severity. Similarly, for mortality outcome, covariate of female gender, proportion of pulmonary disease, diabetes, hypertension, and cardiovascular diseases were significant, these covariates collectively explained R^2^=53% of the between-study variability for mortality.

**Conclusions:** Our findings suggest that obesity is significantly associated with increased severity and higher mortality among COVID-19 patients. Therefore, the inclusion of obesity or its surrogate body mass index in prognostic scores and streamlining the management strategy and treatment guidelines to account for the impact of obesity in patient care management is recommended.

## Introduction

The entire world is enduring the effects of the global coronavirus disease 2019 (COVID-19) pandemic since its inception in December 2019 when pneumonia of unknown origin was diagnosed in Hubei province, Wuhan, China^1, 2^. It was later in January 2020 that the novel coronavirus strand was isolated and subsequently named severe acute respiratory syndrome coronavirus 2 (SARS-CoV-2) in February 2020^3, 4^. As of now, till 4^th^ April, 2021, the Covid-19 pandemic has affected 131,129,824 individuals and has led to 2,850,174 global deaths^5^. Despite the fact that many treatments have been proposed to combat COVID-19, there is currently no uniformly successful therapy^6–12^. Although it is a widespread disease affecting multiple systems, obesity has been identified as one of the major comorbid factors in patients suffering from COVID-19^13–22^.

Overweight (BMI 25 kg/m^2^-29.9 kg/m^2^) and obesity (BMI 30 kg/m^2^ or more) are a major public health problem, especially during the COVID-19 pandemic, because of their association with increased morbidity and mortality^23, 24^. Berrington de Gonzalez et al. (2010) studied the association of overweight and obesity on overall mortality in 1.46 million white adults over a median follow-up period of 10 years. They found approximately linear relationship in the hazard ratios for the BMI. The hazard ratio for every 5-unit increment of BMI was 1.31 in the BMI range of 25 kg/m^2^ to 49.9 kg/m^2^.^25^. According to the 2017-2018 National Health and Nutrition Examination Survey (NHANES), about 42.5% of U.S. adults aged 20 or more are obese and approximately 9% have class 3 obesity or severe obesity (BMI 40 kg/m^2^or more) ^26^. The prevalence of obesity has been increasing rapidly in the last decade.

According to WHO, the prevalence of obesity has nearly tripled in the last four decades amounting to 13% of the entire world’s adult population^27^. This exponential rise in the obesity rates in the midst of the pandemic is a cause for concern. The interplay between obesity and diabetes mellitus, cardiovascular disease, stroke, dyslipidemia, influenza has been established for a long time. The presence of these comorbid determinants has been related to increased predisposition and severity of COVID-19^28–31^. Many studies have reported increased rates of hospitalization, mechanical ventilation, and mortality in patients with higher BMI^32–36^.

To mitigate the impact of heightened morbidity and mortality associated with COVID-19 infection in patients with obesity, it is vital to be cognizant of the implications of increased BMI and its dynamic interaction with other comorbid components. Hence, we evaluated obesity as a paramount risk factor for mortality and severity in COVID-19 infection, independent of potential confounders via systematic review and meta-regression.

## Methods

### Search method and Strategy

For documentation, we adopted the Preferred Systematic Analyses and Meta-Analysis Reporting Items recommendations^37^. A systematic search was conducted from COVID-19 databases from the pandemic inception through January 31^st^, 2021 for full-length articles focusing on the association of increased BMI/ Obesity in COVID-19 using a pre-specified data extraction protocol including bibliographic information (year of publication, first author), study information (country, sample size), patient characteristics (age, baseline comorbidities, gender), treatment information and outcome data. The search strategy consisted of keywords “SARS-CoV-2”, “COVID-19”, “CORONAVIRUS”, “OBESITY”, “BMI”, “OVERWEIGHT” across the COVID-19 database which included articles from Medline (PubMed), Embase, Science Web, and Cochrane Central Controlled Trials Registry. Studies were included from all over the world, there were no language barriers. Other literature sources such as the BioRxiv (preprints), MedRxiv (preprints), ChemRxiv (preprints), and SSRN (preprints) were searched as well. After following a thorough search, full-length articles meeting the inclusion criteria were evaluated. In an attempt to discover further eligible studies, we manually searched the reference lists of the included studies, and previously published meta-analysis, systematic review, and the relevant literature. We also scanned the clinicaltrials.gov registry for completed, as well as in-progress randomized controlled trials (RCTs).

### Eligibility Criteria

The inclusion criteria for the systematic review are as follows:

1. Studies reporting outcomes such as severity or mortality events, at least one functional endpoint of COVID-19 hospitalized patients with increased BMI.
2. Full text, peer-reviewed articles (Case-studies and case series, randomized controlled trials) were included.

### Study selection

The authors (HK and SSR) downloaded all articles from electronic search to EndNote X9^38^ and duplicates were eliminated. Based on the preset eligibility criteria, each study was reviewed by two reviewers (AT, GSS, HK, NJ, RS, SK and SSR) independently, and disagreements were discussed amongst all author-reviewers and resolved via a consensus. The cases included obese Covid-19 positive hospitalized patients and the controls included the non-obese Covid-19 positive hospitalized patients. Unadjusted and adjusted impact measurements were also extracted where appropriate. From each study, various details including first author name, study type, hospitalized total covid-19 positive patients, the definition of COVID-19 severity, definition of obesity, total obese & non-obese COVID-19 positive patients, patients with high severity and mortality, median age, gender (female sex proportion), hypertension proportion, pulmonary disease proportion, cardiovascular disease proportion, diabetes proportion, dyslipidemia proportion, liver disease proportion were mentioned in a tabulated format in excel sheet. These details are exhibited in Table 1. The included data was checked for accuracy by all authors.

**Table 1:** Study characteristics

| Study | Study Design | Definition of Severity | Total COVID-19 Positive Patients | Total Patients with Obesity | Median Age | Female Sex Proportion | Diabetes Proportion | Heart Disease Proportion | Pulmonary Disease Proportion | Hypertension Proportion |
| --- | --- | --- | --- | --- | --- | --- | --- | --- | --- | --- |
| *Al Heialy et al.*<sup>42</sup> | Cohort | ICU admissions | 286 | 102 | 46.9 | 27.2 | 26.2 | 3.8 | 3.8 | 25.8 |
| *Al-Sabah et al.*<sup>43</sup> | Cohort | ICU admissions | 1158 | 157 | 40.5 | 18.4 | 23.4 | N/A | N/A | 20.4 |
| *Alkhatib et al.*<sup>44</sup> | Retrospective cohort | ICU admissions | 158 | 96 | 57 | 61.4 | 48.1 | 13.3 | 17.5 | 67.7 |
| *Anderson et al.*<sup>45</sup> | Retrospective cohort | Intubation | 2466 | 785 | 67 | 42 | 40 | 7 | 17 | 52 |
| *Andrea Rossi et al.*<sup>116</sup> | Cohort | Not valid | 95 | 35 | N/A | 18 | 19 | 38.9 | N/A | 47.4 |
| *Argenziano et al.*<sup>107</sup> | Cohort | ICU admission | 1000 | 352 | 63 | 40.4 | 37.2 | 23.3 | 22.3 | 60.1 |
| *Arjun S et al.*<sup>46</sup> | Cohort | ICU admissions | 142 | 54 | N/A | N/A | N/A | N/A | N/A | N/A |
| *Bellan et al.*<sup>115</sup> | Cohort | Not valid | 407 | 60 | 71 | 41 | 24 | 30 | 3 | 58 |
| *Bhatraju et al.*<sup>47</sup> | Cohort | ICU admissions | 24 | 13 | 64 | 38 | 58 | N/A | 16 | N/A |
| *Biscarin et al.*<sup>48</sup> | Retrospective cohort | ICU admissions | 427 | 80 | 67 | 31.8 | 19 | 28 | N/A | 50 |
| *Borobia et al.*<sup>140</sup> | Cohort | Not valid | 2226 | 242 | 61 | 51.8 | 17.1 | 19.3 | 13.3 | 41.3 |
| *Burrell et al.*<sup>49</sup> | Cohort | Mechanical ventilation | 204 | 80 | 63.5 | 31 | 28 | 20 | 11 | 24 |
| *Busetto et al.*<sup>50</sup> | Cohort | ICU admission | 92 | 29 | 70.5 | 39.1 | 30.4 | 31.5 | 13 | 64.1 |
| *Cai et al.*<sup>51</sup> | Cohort | ICU admission | 383 | 41 | 48 | 52.2 | 7.3 | 4.9 | N/A | 15.1 |
| *Cariou et al.*<sup>52</sup> | Cohort | Mechanical ventilation | 1117 | 428 | 69.8 | 35.1 | 88.5 | 11.6 | 10.4 | 77.2 |
| *Carrillo-Vega et al.*<sup>117</sup> | Cohort | Not valid | 3922 | 987 | 54.2 | 35 | 30 | 5.1 | 8.1 | 34.1 |
| *Castelnuova et al.*<sup>133</sup> | Cohort | Not valid | 3894 | 376 | 67 | 38.3 | 19 | 21.1 | 14.3 | 49.4 |
| *caussy et al.*<sup>53</sup> | Cohort | Mechanical ventilation | 291 | 96 | N/A | N/A | N/A | N/A | N/A | N/A |
| *Cedano et al.*<sup>118</sup> | Cohort | Not valid | 132 | 59 | 63 | 41 | 45 | 24 | 13 | 59 |
| *Chand et al.*<sup>111</sup> | Cohort | Not valid | 300 | 163 | 58.2 | 39.3 | 44.7 | 13.7 | 18.7 | 66.7 |
| *Chao et al.*<sup>54</sup> | Cohort | ICU admissions | 46 | 12 | 13.1 | 32.7 | N/A | 2.1 | 24 | N/A |
| *Ciceri et al.*<sup>112</sup> | Prospective case control | Not valid | 410 | 78 | 65 | 27.1 | 17.8 | 12.4 | 5.3 | 49.5 |
| *Claudia et al.*<sup>99</sup> | Cohort | ICU admissions | 99 | 27 | 67 | 37 | 22 | 28 | 21 | 57 |

**Table 1:** Study characteristics (continued)
|  |  |  |  |  |  |  |  |  |  |  |
| --- | --- | --- | --- | --- | --- | --- | --- | --- | --- | --- |
| **Cravedi *et al.*<sup>113</sup>** | Cohort | Not valid | 144 | 71 | 62 | 34.02 | 52 | 28 | 18.8 | 95 |
| **Czernichow *et al.*<sup>55</sup>** | Cohort | ICU admissions | 5,795 | 1264 | 58 | 34.5 | 42.6 | 4.5 | N/A | 53.4 |
| **de Andrade *et al.*<sup>114</sup>** | Cohort | Not valid | 89405 | 655 | 58.9 | 43.5 | 1.5 | N/A | N/A | 4.2 |
| **Docherty *et al.*<sup>119</sup>** | Cohort | Not valid | 20133 | 1685 | 72.9 | 40.1 | 20.7 | 30.9 | 16.11 | N/A |
| **Dreher *et al.*<sup>56</sup>** | Cohort | ARDS | 50 | 17 | 65 | 34 | 58 | N/A | 50 | 70 |
| **Ebinger *et al.*<sup>57</sup>** | Cohort | ICU admissions | 214 | 44 | 62 | 37 | 40 | 21 | 20 | 54.6 |
| **Fava *et al.*<sup>134</sup>** | Cohort | Not valid | 104 | 28 | 59.7 | 42.3 | 30.8 | 29.8 | 15.4 | 86.5 |
| **Feuth *et al.*<sup>58</sup>** | Cohort | ICU admissions | 28 | 10 | 56 | 46 | 25 | N/A | 21 | 43 |
| **Fusco *et al.*<sup>100</sup>** | Cohort | ICU admission +<br>Mechanical ventilation | 173942 | 46965 | 63 | 48.9 | 40.7 | 73.5 | 22.2 | 64.8 |
| **Gao *et al.*<sup>59</sup>** | Cohort | Not mentioned | 150 | 75 | 48 | 37.8 | 19.3 | N/A | N/A | N/A |
| **Genny Carrillo *et al.*<sup>101</sup>** | Retrospective cohort | Mechanical ventilation | 69334 | 16272 | 55.29 | 37.38 | 30.92 | 4.21 | 5.65 | 34.41 |
| **Gerotziafas *et al.*<sup>60</sup>** | Cohort | ICU admissions | 430 | 67 | 64.3 | 61 | 21.6 | N/A | 9 | 47.7 |
| **Giacomelli *et al.*<sup>121</sup>** | Cohort | Not valid | 233 | 38 | 61 | 30.9 | N/A | N/A | N/A | N/A |
| **Giorgi Rossi *et al.*<sup>122</sup>** | Cohort | Not valid | 1075 | 34 | 63.2 | 38.8 | 17 | 31 | 8.4 | 26 |
| **Goyal *et al.*<sup>61</sup>** | Cohort | Mechanical ventilation | 393 | 136 | 62.2 | 39.4 | 25.2 | 13.7 | 17.6 | 50.1 |
| **Guner *et al.*<sup>62</sup>** | Cohort | ARDS, sepsis, and<br>septic shock | 222 | 77 | 50.6 | 40.5 | 13.5 | 23.6 | 5.4 | 23.4 |
| **Hajifathalian *et al.*<sup>63</sup>** | Cohort | ICU admissions | 770 | 277 | 64 | 39.2 | 31 | 21 | 17.4 | 56.1 |
| **Halasz *et al.*<sup>123</sup>** | Cohort | Not valid | 242 | 48 | 64 | 18.2 | 15.3 | 14.5 | 8.7 | 45.5 |
| **Halvatsiotis *et al.*<sup>139</sup>** | Cohort | Not valid | 90 | 31 | 65.5 | 20 | 18.8 | 21.1 | 12 | 50 |
| **Hojo de Souza *et al.*<sup>120</sup>** | Retrospective cohort | Not valid | 44128 | 3633 | N/A | 45.85 | 39.82 | 52.02 | 6.31 | N/A |
| **Hsu *et al.*<sup>108</sup>** | Cohort | ICU admissions | 1088 | 565 | 63 | 45 | 35.7 | 19.7 | 24.3 | 57 |
| **Hur *et al.*<sup>64</sup>** | Cohort | Intubation | 486 | 259 | 59 | 44.2 | 32.9 | 22.8 | 16 | 54.9 |
| **Ioannou *et al.*<sup>66</sup>** | Cohort | Mechanical ventilation | 3465 | 1412 | 61.1 | 19.6 | 48.8 | 47.3 | 33 | 75 |
| **John Xie *et al.*<sup>106</sup>** | Retrospective cohort | ICU admissions | 287 | 187 | 61.5 | 56.8 | 53.6 | 14.3 | 20.5 | 80.1 |
| **Kaeuffer *et al.*<sup>67</sup>** | Cohort | Death or admission to<br>ICU | 1045 | 351 | 66.3 | 41.4 | 25.3 | 11.6 | 16.5 | 52.4 |
| **Kalligeros *et al.*<sup>32</sup>** | Cohort | ICU admissions | 103 | 49 | 60 | 39.8 | 36.8 | 24.2 | 19.5 | 64 |
| **Kates *et al.*<sup>124</sup>** | Cohort | Not valid | 482 | 166 | 57.5 | 28.8 | 51 | 30.1 | 10.4 | 77.4 |
| **Klang *et al.*** | Cohort | Not valid | 3406 | 1231 | N/A | N/A | N/A | N/A | N/A | N/A |
| **Laccarino *et al.*<sup>65</sup>** | Cohort | ICU admissions | 2378 | 157 | 68.2 | 37.4 | 18.2 | 26.4 | 8.5 | 58.5 |

**Table 1:** Study characteristics (continued)
|  |  |  |  |  |  |  |  |  |  |  |
| --- | --- | --- | --- | --- | --- | --- | --- | --- | --- | --- |
| **Lighter *et al.*<sup>68</sup>** | Cohort | ICU admissions | 3615 | 1370 | N/A | N/A | N/A | N/A | N/A | N/A |
| **Ling Hu *et al.*<sup>102</sup>** | Cohort | Not mentioned | 323 | 13 | 61 | 48.6 | 14.6 | 12.7 | 10.9 | 32.5 |
| **Lodigiani *et al.*<sup>69</sup>** | Cohort | ICU admissions | 361 | 87 | 66 | 32 | 22.7 | N/A | N/A | 47.2 |
| **Marcello *et al.*<sup>135</sup>** | Cohort | Not valid | 6248 | 2278 | 61 | 38 | 33 | 31 | 11 | 37 |
| **Mejía-Vilet.<sup>70</sup>** | Cohort | ICU admissions | 329 | 132 | 49 | 36 | 24 | N/A | N/A | 27 |
| **Mendy *et al.*<sup>71</sup>** | Cohort | Death or admission to ICU | 216 | 53 | 60 | 44.4 | 43.5 | 77.3 | 16.7 | N/A |
| **Menezes Soares *et al.*<sup>136</sup>** | Cohort | Not valid | 1152 | 111 | N/A | 42.8 | 24 | 45.6 | 9.6 | N/A |
| **Mikami *et al.*<sup>137</sup>** | Cohort | Not valid | 3708 | 288 | 66 | 43 | 24.3 | N/A | 8.7 | 34.3 |
| **Monteiro *et al.*<sup>72</sup>** | Cohort | Mechanical ventilation | 112 | 40 | 61 | 34 | 64 | 15 | 17.4 | 50 |
| **Motaib *et al.*<sup>169</sup>** | Cohort | ICU admissions | 107 | 24 | 53 | 40 | 15 | 15 | 8.4 | 30.8 |
| **Mughal *et al.*<sup>73</sup>** | Cohort | Mechanical ventilation | 129 | 18 | 63 | 37.2 | 19.4 | 17.1 | 10.9 | 43.3 |
| **Murillo-Zamora *et al.*<sup>126</sup>** | Cohort | Not valid | 5393 | 1197 | N/A | 36.4 | 31.1 | N/A | 7.8 | 36.6 |
| **Nachega *et al.*<sup>74</sup>** | Cohort | ICU admissions | 766 | 39 | 46 | 34.4 | 14 | 3.9 | 3.4 | 25.4 |
| **Nakeshbandi *et al.*<sup>75</sup>** | Cohort | Mechanical ventilation | 504 | 215 | 68 | 48 | 53 | 19 | 16 | 83 |
| **Newton *et al.*<sup>76</sup>** | Cohort | Death or admission to ICU | 370 | 102 | 62.2 | 51.8 | 42.3 | 17.4 | 2.5 | 66.8 |
| **Olivas-Martínez *et al.*<sup>127</sup>** | Cohort | Not valid | 800 | 357 | 51.9 | 39 | 26 | 4.6 | 2.3 | 30 |
| **Ortiz-Brizuela *et al.*<sup>77</sup>** | Cohort | ICU admissions | 140 | 50 | 49 | 29.3 | 22.9 | 4.3 | 2.8 | 32 |
| **Palaiodimos *et al.*<sup>78</sup>** | Cohort | ICU admissions | 200 | 46 | 64 | 51 | 39.5 | 33.5 | 27.5 | 76 |
| **Parker *et al.*<sup>128</sup>** | Cohort | Not valid | 113 | 32 | 48 | 61 | 39 | 5.4 | 17.2 | 42 |
| **Parra-Bracamonte *et al.*<sup>129</sup>** | Cohort | Not valid | 95458 | 22390 | 44 | 38.1 | 31.3 | 4.2 | 5.8 | 35.1 |
| **Peng *et al.*<sup>138</sup>** | Cohort | Not mentioned | 112 | 33 | 62 | 59 | N/A | 55.3 | N/A | 82 |
| **Pepe *et al.*<sup>79</sup>** | Cohort | ICU admission | 2214 | 440 | 49.6 | 40.2 | 9.1 | 8 | 12.3 | 26.2 |
| **Petersen, A. *et al.*<sup>80</sup>** | Cohort | ICU admissions | 30 | 19 | 65.6 | 12 | 5 | 5 | 3 | 15 |
| **Petrilli *et al.*<sup>81</sup>** | Cohort | Mechanical ventilation and ICU admission | 2741 | 1081 | 63 | 38.8 | 34.7 | 34.8 | 16.5 | 62 |
| **Pettit *et al.*<sup>82</sup>** | Cohort | ICU admissions | 238 | 146 | 58.5 | 52.5 | 28.6 | 21.4 | 26.5 | 52.9 |
| **Philipose *et al.*<sup>83</sup>** | Cohort | Not mentioned | 368 | 125 | 72 | 40.5 | 35.4 | 33.6 | 28.1 | 50.2 |
| **Pongpirul *et al.*<sup>84</sup>** | Cohort | Not mentioned | 193 | 22 | 37 | 41.5 | 8.3 | 1 | 1.6 | 16.1 |
| **Rachel *et al.*<sup>103</sup>** | Cohort | Mechanical ventilation | 305 | 127 | 60 | 42 | 35 | 21 | 18 | 52 |

**Table 1:** Study characteristics (continued)
|  |  |  |  |  |  |  |  |  |  |  |
| --- | --- | --- | --- | --- | --- | --- | --- | --- | --- | --- |
| **Ramlall *et al.*<sup>85</sup>** | Cohort | Mechanical ventilation | 6393 | 831 | 57.1 | 50.3 | 14.2 | 26.6 | N/A | 31.1 |
| **Rao *et al.*<sup>86</sup>** | Cohort | Not mentioned | 240 | 114 | 48 | 53.8 | 9.6 | 17.9 | 1.2 | N/A |
| **Reilev *et al.*<sup>87</sup>** | Cohort | ICU admissions | 450 | 50 | 81 | 38 | 26 | 45 | 28 | 74 |
| **Rodriguez *et al.*<sup>130</sup>** | Cohort | Not valid | 43 | 11 | 65.5 | 27.2 | 18.6 | 14 | 9.3 | 30.2 |
| **Rodríguez-Molinero *et al.*<sup>88</sup>** | Cohort | Need for oxygen therapy via nonrebreather mask or mechanical ventilation | 418 | 74 | N/A | 57 | 23.7 | 24.1 | N/A | 52 |
| **Rottoli *et al.*<sup>89</sup>** | Cohort | ICU admissions | 482 | 104 | 66.2 | 37.3 | 15.2 | 21.2 | 13.1 | 51.7 |
| **Salacup G *et al.*<sup>131</sup>** | Cohort | Not valid | 242 | 97 | 66 | 49.17 | 49 | 14.8 | 19.8 | 74 |
| **Shah *et al.*<sup>132</sup>** | Cohort | Not valid | 522 | 481 | 63 | 58.2 | 42.3 | 22.6 | 22 | 79.7 |
| **Shekhar *et al.*<sup>90</sup>** | Cohort | ICU admissions | 50 | 20 | 55.5 | 54 | 36 | 14 | 16 | 34 |
| **Simmonet *et al.*<sup>152</sup>** | Cohort | Mechanical ventilation | 124 | 59 | 60 | 27 | 23 | N/A | N/A | 49 |
| **Steinberg *et al.*<sup>92</sup>** | Cohort | Mechanical ventilation | 210 | 100 | N/A | N/A | N/A | N/A | N/A | N/A |
| **Suleyman *et al.*<sup>93</sup>** | Cohort | ICU admission | 355 | 210 | 61.4 | 53.5 | 43.4 | 29.1 | 40.3 | 72.7 |
| **Tonetti *et al.*<sup>94</sup>** | Cohort | ICU admissions | 700 | 176 | 69.4 | 23 | 21.8 | 61.8 | 14.4 | N/A |
| **Urra *et al.*<sup>95</sup>** | Retrospective case control | ICU admissions | 172 | 17 | N/A | 39.5 | 22.6 | 16.27 | 9.8 | 50.5 |
| **Vaquero-Roncero *et al.*<sup>96</sup>** | Cohort | ICU admissions | 146 | 46 | N/A | N/A | N/A | N/A | N/A | N/A |
| **Wang J *et al.*<sup>104</sup>** | Cohort | ICU admissions | 297 | 40 | 44.3 | 44.7 | 8.41 | 2.02 | 4.04 | 16.16 |
| **Wang Min *et al.*<sup>3</sup>** | Retrospective cohort | Respiratory failure | 541 | 60 | 52 | 45.29 | 8.69 | 5.36 | N/A | 24.77 |
| **Wang R *et al.*<sup>97</sup>** | Cohort | Not mentioned | 96 | 44 | N/A | N/A | N/A | N/A | N/A | 52.1 |
| **Xiang Ong *et al.*<sup>105</sup>** | Cohort | ICU admissions | 91 | 40 | 55 | 44 | 19.7 | 9.9 | N/A | 33 |
| **Zheng *et al.*<sup>98</sup>** | Cohort | Not mentioned | 66 | 45 | 47 | 74.2 | 24.2 | N/A | N/A | 28.8 |

Reporting Items for Systematic Reviews and Meta-Analysis (PRISMA) guidelines were used. Figure 1.

**Figure 1:**
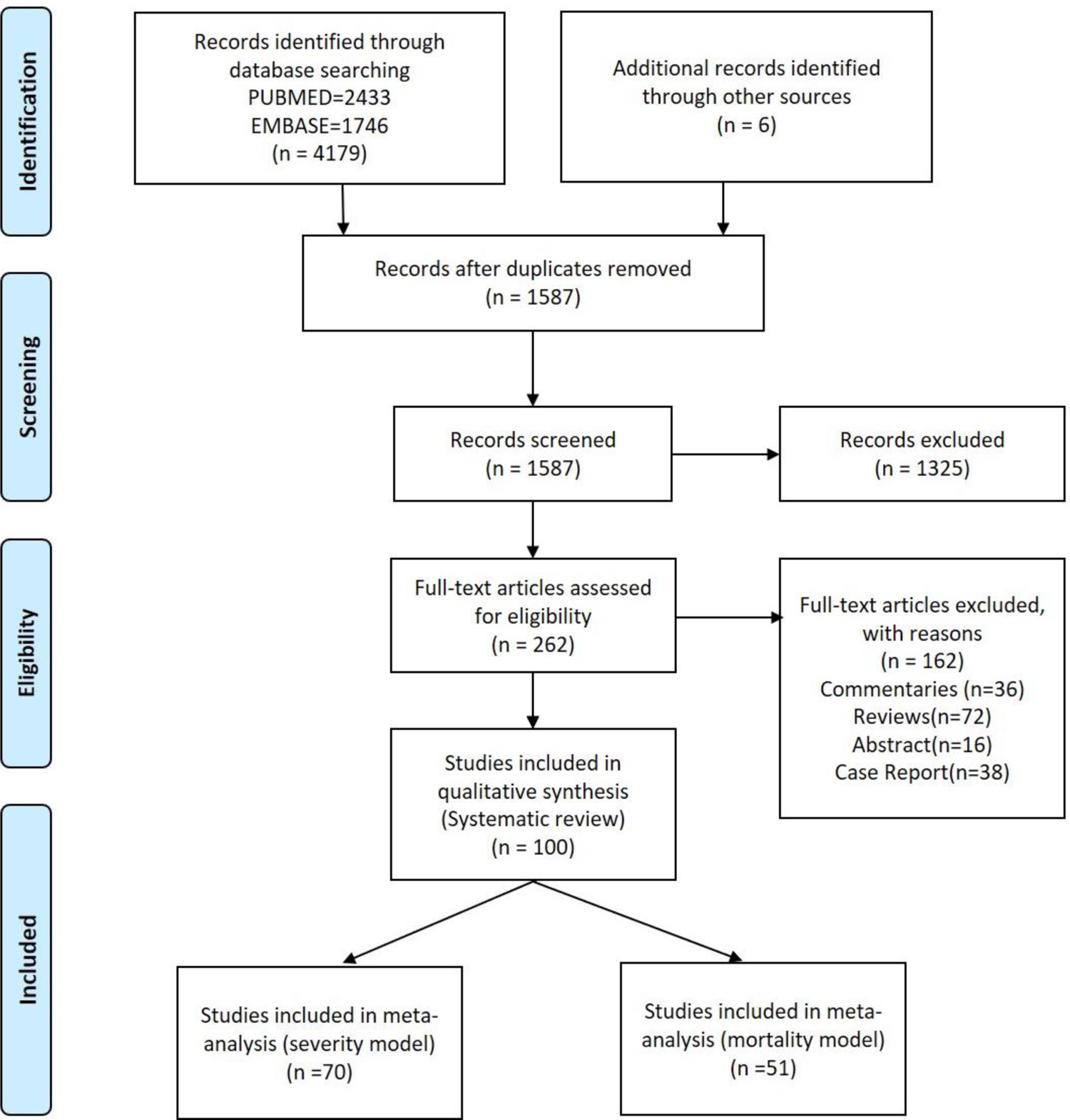
Prisma flow diagram

### Outcomes

All-cause severity in hospitalized COVID-19 patients with high BMI/ Obesity was the primary outcome. The severity rate was evaluated in comparison to the control group (non-obese COVID-19 hospitalized patients). While all-cause mortality in COVID-19 hospitalized patients with high BMI/ obesity was the secondary outcome.

### Statistical analysis

The meta-analysis specifically included case-control and cohort studies comparing the effects of high BMI/Obesity in COVID-19 hospitalized patients comparing them to the non-obese COVID-19 hospitalized patients. All outcomes were analyzed using the Mantel-Haenszel method for dichotomous data to estimate pooled risk ratio (RR) utilizing the Review Manager (RevMan)-Version 5.4, The Cochrane Collaboration, 2020. Meta-analysis was performed first for studies reporting severity of patients in both groups followed by that for studies reporting severity of disease assuming independence of results for studies that reported both. Due to anticipated heterogeneity, summary statistics were calculated using a random-effects model. This model accounts for variability between studies as well as within studies. Statistical heterogeneity was assessed using Q value and I^2^ statistics.

To explore differences between studies that might be expected to influence the effect size, we performed random effects (maximum likelihood method) univariate and multivariate meta-regression analyses. The potential sources of variability hypothesized were the gender of the study sample, the proportion of subjects with diabetes, pulmonary disease, cardiovascular disease, and hypertension. Covariates were selected for further modeling if they significantly (P < 0.05) modified the association between mortality or severity in the COVID-19 hospitalized patients with high BMI/Obesity. Two models were created, one for severity and the other for mortality of disease as outcomes. Subsequently, preselected covariates were included in a manual backward and stepwise multiple meta-regression analysis with P = 0.05 as a cutoff point for removal. P < 0.05. (P < 0.10 for heterogeneity) was considered statistically significant. All meta-analysis and meta-regression tests were 2-tailed. The meta-regression was done with the Comprehensive Meta-Analysis software package (Biostat, Englewood, NJ, USA)14^39^.

We conducted sensitivity analysis with BMI categories (BMI <18 kg/m^2^, BMI 18 kg/m^2^-25 kg/m^2^, BMI 25 kg/m^2^-29.9 kg/m^2^, BMI >30 kg/m^2^, and BMI>40 kg/m^2^) to decrease inherent selection bias in observational studies^40^.

### Risk of Bias

Risk of Bias assessment-The Newcastle-Ottawa (NOS) scale12 was used for measuring the risk of bias in case-control studies and cohort studies. The following classes were rated per study: low bias risk (9 points), moderate bias risk (5-7 points), and high bias risk (0-4 items. For a cross-sectional study, we used the modified version of NOS, assigning the study in the following groups: Low risk of bias (8-10), moderate risk (5-7), high risk of bias (0-4))^41^. Three reviewers (AT, SA, and SSR) evaluated the likelihood of bias independently, and any conflict was resolved by consensus (Table 2A and 2B).

**Table 2:** Quality Assessment Tool for Observational Cohort and Cross-Sectional Studies, **2a**: Cohort studies, **2b:** Case control studies

| Criteria | Al Heialy *et al.*<sup>42</sup> | Al-Sabah *et al.*<sup>43</sup> | Alkhatab *et al.*<sup>44</sup> | Anderson *et al.*<sup>45</sup> | Andrea Rossi *et al.*<sup>116</sup> | Argenzi *et al.*<sup>107</sup> | Arjun S *et al.*<sup>46</sup> | Bellani *et al.*<sup>115</sup> |
| --- | --- | --- | --- | --- | --- | --- | --- | --- |
| Was the research question or objective in this paper clearly stated? | Yes | Yes | Yes | Yes | Yes | Yes | Yes | Yes |
| Was the study population clearly specified and defined? | Yes | Yes | Yes | Yes | Yes | Yes | Yes | Yes |
| Was the participation rate of eligible persons at least 50%? | Yes | Yes | Yes | Yes | Yes | Yes | No | Yes |
| Were all the subjects selected or recruited from the same or similar populations (including the same time period)? Were inclusion and exclusion criteria for being in the study prespecified and applied uniformly to all participants? | Yes | Yes | Yes | Yes | Yes | Yes | Yes | Yes |
| Was a sample size justification, power description, or variance and effect estimates provided? | Yes | Yes | Yes | Yes | Yes | Yes | No | Yes |
| For the analyses in this paper, were the exposure(s) of interest measured prior to the outcome(s) being measured? | Yes | Yes | Yes | Yes | Yes | Yes | Yes | Yes |
| Was the timeframe sufficient so that one could reasonably expect to see an association between exposure and outcome if it existed? | No | No | No | No | No | No | No | No |
| For exposures that can vary in amount or level, did the study examine different levels of the exposure as related to the outcome (e.g., categories of exposure, or exposure measured as continuous variable)? | Yes | Yes | No | Yes | No | No | Yes | NA |
| Were the exposure measures (independent variables) clearly defined, valid, reliable, and implemented consistently across all study participants? | Yes | Yes | No | Yes | Yes | Yes | Yes | Yes |
| Was the exposure(s) assessed more than once over time? | NA | NA | No | No | No | No | No | No |
| Were the outcome measures (dependent variables) clearly defined, valid, reliable, and implemented consistently across all study participants? | Yes | Yes | No | Yes | Yes | NA | NA | NA |
| Were the outcome assessors blinded to the exposure status of participants? | CD | CD | CD | CD | CD | Yes | NA | Yes |
| Was loss to follow-up after baseline 20% or less? | Yes | Yes | Yes | Yes | Yes | Yes | Yes | Yes |
| Were key potential confounding variables measured and adjusted statistically for their impact on the relationship between exposure(s) and outcome(s)? | Yes | Yes | Yes | Yes | Yes | Yes | No | Yes |
| **Quality Rating (Good, Fair, or Poor)** | Good | Good | Low | Good | Good | Good | Low | Good |
| **Risk of Bias** | Low | Low | Moderate | Low | Low | Low | Moderate | Low |

CD, cannot determine; NA, not applicable; NR, not reported

| Criteria | Bhatraju *et al.*<sup>47</sup> | Biscarini *et al.*<sup>48</sup> | Borobia *et al.*<sup>140</sup> | Burrell *et al.*<sup>49</sup> | Busetto *et al.*<sup>50</sup> | Cai *et al.*<sup>51</sup> | Cariou *et al.*<sup>52</sup> | Cravedi *et al.*<sup>113</sup> |
| --- | --- | --- | --- | --- | --- | --- | --- | --- |
| Was the research question or objective in this paper clearly stated? | Yes | Yes | Yes | Yes | Yes | Yes | Yes | Yes |
| Was the study population clearly specified and defined? | Yes | Yes | Yes | Yes | Yes | Yes | Yes | Yes |
| Was the participation rate of eligible persons at least 50%? | Yes | Yes | Yes | Yes | Yes | Yes | Yes | Yes |
| Were all the subjects selected or recruited from the same or similar populations (including the same time period)? Were inclusion and exclusion criteria for being in the study prespecified and applied uniformly to all participants? | Yes | Yes | Yes | Yes | Yes | Yes | Yes | Yes |
| Was a sample size justification, power description, or variance and effect estimates provided? | No | Yes | Yes | Yes | No | Yes | Yes | Yes |
| For the analyses in this paper, were the exposure(s) of interest measured prior to the outcome(s) being measured? | No | Yes | Yes | Yes | No | Yes | Yes | Yes |
| Was the timeframe sufficient so that one could reasonably expect to see an association between exposure and outcome if it existed? | NA | No | NO | No | No | No | NA | No |
| For exposures that can vary in amount or level, did the study examine different levels of the exposure as related to the outcome (e.g., categories of exposure, or exposure measured as continuous variable)? | NA | Yes | Yes | No | Yes | NA | CD | Yes |
| Were the exposure measures (independent variables) clearly defined, valid, reliable, and implemented consistently across all study participants? | Yes | Yes | Yes | Yes | Yes | Yes | Yes | Yes |
| Was the exposure(s) assessed more than once over time? | Yes | Yes | Yes | Yes | Yes | Yes | Yes | No |
| Were the outcome measures (dependent variables) clearly defined, valid, reliable, and implemented consistently across all study participants? | NA | NA | Na | NA | Yes | NA | Yes | Yes |
| Were the outcome assessors blinded to the exposure status of participants? | NA | Yes | Yes | Yes | NA | NA | CD | CD |
| Was loss to follow-up after baseline 20% or less? | Yes | Yes | Yes | Yes | NA | Yes | Yes | Yes |
| Were key potential confounding variables measured and adjusted statistically for their impact on the relationship between exposure(s) and outcome(s)? | CD | Yes | Yes | No | Yes | Yes | Yes | Yes |
| **Quality Rating (Good, Fair, or Poor)** | Low | High | High | Good | Low | Good | Good | Good |
| **Risk of Bias** | Moderate | Very low | Very low | Low | Moderate | Low | Low | Low |

CD, cannot determine; NA, not applicable; NR, not reported

| Criteria | Carrillo-Vega *et al.*<sup>117</sup> | Castelnuova *et al.*<sup>133</sup> | Caussy *et al.*<sup>53</sup> | Cedano *et al.*<sup>118</sup> | Chand *et al.*<sup>111</sup> | Chao *et al.*<sup>54</sup> | Claudia *et al.*<sup>99</sup> | Czernichow *et al.*<sup>55</sup> |
| --- | --- | --- | --- | --- | --- | --- | --- | --- |
| Was the research question or objective in this paper clearly stated? | Yes | Yes | Yes | Yes | Yes | Yes | Yes | Yes |
| Was the study population clearly specified and defined? | Yes | Yes | Yes | Yes | Yes | Yes | Yes | Yes |
| Was the participation rate of eligible persons at least 50%? | Yes | Yes | yes | Yes | Yes | Yes | Yes | Yes |
| Were all the subjects selected or recruited from the same or similar populations (including the same time period)? Were inclusion and exclusion criteria for being in the study prespecified and applied uniformly to all participants? | Yes | Yes | Yes | Yes | Yes | Yes | Yes | Yes |
| Was a sample size justification, power description, or variance and effect estimates provided? | Yes | Yes | No | Yes | Yes | Yes | Yes | Yes |
| For the analyses in this paper, were the exposure(s) of interest measured prior to the outcome(s) being measured? | Yes | Yes | No | Yes | Yes | Yes | Yes | Yes |
| Was the timeframe sufficient so that one could reasonably expect to see an association between exposure and outcome if it existed? | No | NO | NA | No | No | No | NO | No |
| For exposures that can vary in amount or level, did the study examine different levels of the exposure as related to the outcome (e.g., categories of exposure, or exposure measured as continuous variable)? | No | Yes | CD | Yes | Yes | Yes | Yes | CD |
| Were the exposure measures (independent variables) clearly defined, valid, reliable, and implemented consistently across all study participants? | Yes | Yes | Yes | Yes | Yes | Yes | Yes | Yes |
| Was the exposure(s) assessed more than once over time? | No | Yes | Yes | Yes | Yes | Yes | Yes | Yes |
| Were the outcome measures (dependent variables) clearly defined, valid, reliable, and implemented consistently across all study participants? | Yes | Yes | CD | Yes | NA | Yes | Na | NA |
| Were the outcome assessors blinded to the exposure status of participants? | CD | CD | CD | CD | NA | CD | Yes | CD |
| Was loss to follow-up after baseline 20% or less? | Yes | NA | Yes | NA | Yes | Yes | Yes | Yes |
| Were key potential confounding variables measured and adjusted statistically for their impact on the relationship between exposure(s) and outcome(s)? | Yes | Yes | Yes | Yes | Yes | Yes | Yes | Yes |
| **Quality Rating (Good, Fair, or Poor)** | Good | Good | Low | Good | Good | High | High | Good |
| **Risk of Bias** | Low | Low | Moderate | Low | Low | Very low | Very low | Low |

CD, cannot determine; NA, not applicable; NR, not reported

| Criteria | de Andrade *et al.*<sup>114</sup> | Docherty *et al.*<sup>119</sup> | Dreher *et al.*<sup>56</sup> | Ebinger *et al.*<sup>57</sup> | Fava *et al.*<sup>134</sup> | Feuth *et al.*<sup>58</sup> | Fusco *et al.*<sup>100</sup> | Gao *et al.*<sup>59</sup> |
| --- | --- | --- | --- | --- | --- | --- | --- | --- |
| Was the research question or objective in this paper clearly stated? | Yes | Yes | Yes | Yes | Yes | Yes | Yes | Yes |
| Was the study population clearly specified and defined? | Yes | Yes | Yes | Yes | Yes | Yes | Yes | Yes |
| Was the participation rate of eligible persons at least 50%? | Yes | Yes | Yes | Yes | Yes | Yes | Yes | Yes |
| Were all the subjects selected or recruited from the same or similar populations (including the same time period)? Were inclusion and exclusion criteria for being in the study prespecified and applied uniformly to all participants? | Yes | Yes | Yes | Yes | Yes | Yes | Yes | Yes |
| Was a sample size justification, power description, or variance and effect estimates provided? | Yes | Yes | Yes | Yes | Yes | Yes | Yes | Yes |
| For the analyses in this paper, were the exposure(s) of interest measured prior to the outcome(s) being measured? | No | Yes | Yes | Yes | Yes | Yes | Yes | Yes |
| Was the timeframe sufficient so that one could reasonably expect to see an association between exposure and outcome if it existed? | No | No | No | No | No | No | No | NO |
| For exposures that can vary in amount or level, did the study examine different levels of the exposure as related to the outcome (e.g., categories of exposure, or exposure measured as continuous variable)? | No | Yes | Yes | No | No | Yes | Yes | Yes |
| Were the exposure measures (independent variables) clearly defined, valid, reliable, and implemented consistently across all study participants? | Yes | Yes | No | Yes | No | No | No | No |
| Was the exposure(s) assessed more than once over time? | Yes | Yes | Yes | Yes | Yes | Yes | No | No |
| Were the outcome measures (dependent variables) clearly defined, valid, reliable, and implemented consistently across all study participants? | Yes | Yes | Yes | Yes | Yes | Yes | Yes | Yes |
| Were the outcome assessors blinded to the exposure status of participants? | NR | Yes | CD | CD | Yes | CD | Yes | CD |
| Was loss to follow-up after baseline 20% or less? | Yes | Yes | Yes | Yes | Yes | Yes | Yes | Yes |
| Were key potential confounding variables measured and adjusted statistically for their impact on the relationship between exposure(s) and outcome(s)? | Yes | Yes | Yes | Yes | Yes | Yes | Yes | Yes |
| **Quality Rating (Good, Fair, or Poor)** | Good | High | Good | Good | Good | Good | Good | Good |
| **Risk of Bias** | Low | Very low | Low | Low | Low | Low | Low | Low |

CD, cannot determine; NA, not applicable; NR, not reported

| Criteria | Genny Carrillo *et al.*<sup>101</sup> | Gerotziafas *et al.*<sup>60</sup> | Giacome Ili *et al.*<sup>121</sup> | Giorgi rossi *et al.*<sup>122</sup> | Goyal *et al.*<sup>61</sup> | Guner *et al.*<sup>62</sup> | Hajifath alian *et al.*<sup>63</sup> | Halasz *et al.*<sup>123</sup> |
| --- | --- | --- | --- | --- | --- | --- | --- | --- |
| Was the research question or objective in this paper clearly stated? | Yes | Yes | Yes | Yes | Yes | Yes | Yes | Yes |
| Was the study population clearly specified and defined? | Yes | Yes | yes | Yes | Yes | Yes | Yes | Yes |
| Was the participation rate of eligible persons at least 50%? | Yes | Yes | yes | Yes | Yes | Yes | Yes | Yes |
| Were all the subjects selected or recruited from the same or similar populations (including the same time period)? Were inclusion and exclusion criteria for being in the study prespecified and applied uniformly to all participants? | Yes | Yes | Yes | Yes | Yes | Yes | Yes | Yes |
| Was a sample size justification, power description, or variance and effect estimates provided? | Yes | Yes | Yes | Yes | No | Yes | Yes | Yes |
| For the analyses in this paper, were the exposure(s) of interest measured prior to the outcome(s) being measured? | Yes | Yes | Yes | Yes | NO | Yes | Yes | Yes |
| Was the timeframe sufficient so that one could reasonably expect to see an association between exposure and outcome if it existed? | No | Yes | Yes | Yes | No | No | No | No |
| For exposures that can vary in amount or level, did the study examine different levels of the exposure as related to the outcome (e.g., categories of exposure, or exposure measured as continuous variable)? | Yes | No | No | No | No | Yes | Yes | Yes |
| Were the exposure measures (independent variables) clearly defined, valid, reliable, and implemented consistently across all study participants? | NO | Yes | No | No | Yes | No | Yes | Yes |
| Was the exposure(s) assessed more than once over time? | No | Yes | Yes | No | No | No | Yes | NO |
| Were the outcome measures (dependent variables) clearly defined, valid, reliable, and implemented consistently across all study participants? | Yes | Yes | Yes | Yes | Yes | Yes | Yes | Yes |
| Were the outcome assessors blinded to the exposure status of participants? | CD | CD | No | CD | No | CD | CD | CD |
| Was loss to follow-up after baseline 20% or less? | Yes | Yes | Yes | Yes | Yes | Yes | Yes | Yes |
| Were key potential confounding variables measured and adjusted statistically for their impact on the relationship between exposure(s) and outcome(s)? | No | No | No | Yes | No | Yes | Yes | Yes |
| **Quality Rating (Good, Fair, or Poor)** | Fair | Fair | Good | Good | Low | Good | High | Good |
| **Risk of Bias** | Unclear | Unclear | Low | Low | Moderate | Low | Very low | Low |

CD, cannot determine; NA, not applicable; NR, not reported

| Criteria | Halvatsi<br>otis et<br>al.<sup>139</sup> | Hojo de<br>Souza et<br>al.<sup>120</sup> | Hsu et<br>al.<sup>108</sup> | Hur et<br>al.<sup>64</sup> | Laccarin<br>o et al.<sup>65</sup> | Ioannou<br>et al.<sup>66</sup> | John Xie<br>et al.<sup>106</sup> | Kaeuffer<br>et al.<sup>67</sup> |
| --- | --- | --- | --- | --- | --- | --- | --- | --- |
| Was the research question or objective in this paper clearly stated? | Yes | Yes | Yes | Yes | Yes | Yes | Yes | Yes |
| Was the study population clearly specified and defined? | Yes | Yes | Yes | Yes | Yes | Yes | Yes | Yes |
| Was the participation rate of eligible persons at least 50%? | Yes | Yes | Yes | Yes | Yes | Yes | Yes | Yes |
| Were all the subjects selected or recruited from the same or similar populations (including the same time period)? Were inclusion and exclusion criteria for being in the study prespecified and applied uniformly to all participants? | Yes | Yes | Yes | Yes | Yes | Yes | Yes | Yes |
| Was a sample size justification, power description, or variance and effect estimates provided? | Yes | Yes | Yes | Yes | Yes | Yes | Yes | Yes |
| For the analyses in this paper, were the exposure(s) of interest measured prior to the outcome(s) being measured? | Yes | Yes | Yes | Yes | No | Yes | Yes | Yes |
| Was the timeframe sufficient so that one could reasonably expect to see an association between exposure and outcome if it existed? | No | No | No | No | No | No | No | No |
| For exposures that can vary in amount or level, did the study examine different levels of the exposure as related to the outcome (e.g., categories of exposure, or exposure measured as continuous variable)? | Yes | Yes | Yes | Yes | Yes | Yes | Yes | Yes |
| Were the exposure measures (independent variables) clearly defined, valid, reliable, and implemented consistently across all study participants? | Yes | Yes | No | Yes | Yes | Yes | Yes' | Yes |
| Was the exposure(s) assessed more than once over time? | No | No | Yes | No | No | Yes | Yes | Yes |
| Were the outcome measures (dependent variables) clearly defined, valid, reliable, and implemented consistently across all study participants? | Yes | Yes | Yes | Yes | Yes | Yes | Yes | Yes |
| Were the outcome assessors blinded to the exposure status of participants? | CD | CD | CD | CD | Yes | No | Yes | Yes |
| Was loss to follow-up after baseline 20% or less? | Yes | Yes | Yes | Yes | Yes | Yes | Yes | Yes |
| Were key potential confounding variables measured and adjusted statistically for their impact on the relationship between exposure(s) and outcome(s)? | Yes | Yes | No | Yes | NA | Yes | CD | Yes |
| **Quality Rating (Good, Fair, or Poor)** | Good | Good | Good | Good | Good | High | High | High |
| **Risk of Bias** | Low | Low | Low | Low | Low | Very low | Very low | Very low |

CD, cannot determine; NA, not applicable; NR, not reported

| Criteria | Kalligero<br>*s et al.*<sup>32</sup> | Kates *et al.*<sup>124</sup> | Klang *et al.*<sup>125</sup> | Lighter<br>*et al.*<sup>68</sup> | Ling Hu<br>*et al.*<sup>102</sup> | Lodigiani<br>*et al.*<sup>69</sup> | Marcello<br>*et al.*<sup>135</sup> | Mejía-Vilet<br>*et al.*<sup>70</sup> |
| --- | --- | --- | --- | --- | --- | --- | --- | --- |
| Was the research question or objective in this paper clearly stated? | Yes | Yes | Yes | Yes | Yes | Yes | Yes | Yes |
| Was the study population clearly specified and defined? | Yes | Yes | Yes | Yes | Yes | Yes | Yes | Yes |
| Was the participation rate of eligible persons at least 50%? | Yes | Yes | Yes | Yes | Yes | Yes | Yes | Yes |
| Were all the subjects selected or recruited from the same or similar populations (including the same time period)? Were inclusion and exclusion criteria for being in the study prespecified and applied uniformly to all participants? | Yes | Yes | Yes | Yes | Yes | Yes | Yes | Yes |
| Was a sample size justification, power description, or variance and effect estimates provided? | Yes | Yes | Yes | Yes | Yes | Yes | Yes | Yes |
| For the analyses in this paper, were the exposure(s) of interest measured prior to the outcome(s) being measured? | Yes | Yes | Yes | Yes | Yes | Yes | Yes | Yes |
| Was the timeframe sufficient so that one could reasonably expect to see an association between exposure and outcome if it existed? | No | No | No | No | CD | CD | No | No |
| For exposures that can vary in amount or level, did the study examine different levels of the exposure as related to the outcome (e.g., categories of exposure, or exposure measured as continuous variable)? | Yes | Yes | Yes | Yes | Yes | Yes | Yes | Yes |
| Were the exposure measures (independent variables) clearly defined, valid, reliable, and implemented consistently across all study participants? | Yes | Yes | No | No | Yes | Yes | Yes | Yes |
| Was the exposure(s) assessed more than once over time? | Yes | No | Yes | No | No | Yes | No | Yes |
| Were the outcome measures (dependent variables) clearly defined, valid, reliable, and implemented consistently across all study participants? | Yes | Yes | Yes | Yes | Yes | Yes | Yes | Yes |
| Were the outcome assessors blinded to the exposure status of participants? | CD | CD | CD | CD | Yes | Yes | No | No |
| Was loss to follow-up after baseline 20% or less? | Yes | Yes | Yes | Yes | Yes | Yes | Yes | Yes |
| Were key potential confounding variables measured and adjusted statistically for their impact on the relationship between exposure(s) and outcome(s)? | Yes | Yes | Yes | No | Yes | Yes | Yes | Yes |
| **Quality Rating** | High | Good | Good | Fair | High | Good | Good | High |
| **Risk of Bias** | Very low | Low | Low | Unclear | Very low | Low | Low | Very low |

CD, cannot determine; NA, not applicable; NR, not reported

| Criteria | Mendy<br>*et al.*<sup>71</sup> | Menezes<br>Soares<br>*et al.*<sup>136</sup> | Mikami<br>*et al.*<sup>137</sup> | Monteiro<br>*et al.*<sup>72</sup> | Motaib<br>*et al.*<sup>169</sup> | Mughal<br>*et al.*<sup>73</sup> | Murillo-Zamora<br>*et al.*<sup>126</sup> | Nachega<br>*et al.*<sup>74</sup> |
| --- | --- | --- | --- | --- | --- | --- | --- | --- |
| Was the research question or objective in this paper clearly stated? | Yes | Yes | Yes | Yes | Yes | Yes | Yes | Yes |
| Was the study population clearly specified and defined? | Yes | Yes | Yes | Yes | Yes | Yes | Yes | Yes |
| Was the participation rate of eligible persons at least 50%? | Yes | Yes | Yes | Yes | Yes | Yes | Yes | Yes |
| Were all the subjects selected or recruited from the same or similar populations (including the same time period)? Were inclusion and exclusion criteria for being in the study prespecified and applied uniformly to all participants? | Yes | Yes | Yes | Yes | Yes | Yes | Yes | Yes |
| Was a sample size justification, power description, or variance and effect estimates provided? | Yes | Yes | Yes | Yes | Yes | Yes | Yes | Yes |
| For the analyses in this paper, were the exposure(s) of interest measured prior to the outcome(s) being measured? | Yes | Yes | Yes | Yes | Yes | Yes | Yes | Yes |
| Was the timeframe sufficient so that one could reasonably expect to see an association between exposure and outcome if it existed? | Yes | No | No | No | No | Yes | No | No |
| For exposures that can vary in amount or level, did the study examine different levels of the exposure as related to the outcome (e.g., categories of exposure, or exposure measured as continuous variable)? | Yes | Yes | Yes | Yes | Yes | Yes | Yes | Yes |
| Were the exposure measures (independent variables) clearly defined, valid, reliable, and implemented consistently across all study participants? | Yes | Yes | Yes | Yes | Yes | Yes | Yes | Yes |
| Was the exposure(s) assessed more than once over time? | No | Yes | Yes | Yes | No | Yes | Yes | No |
| Were the outcome measures (dependent variables) clearly defined, valid, reliable, and implemented consistently across all study participants? | Yes | Yes | Yes | Yes | Yes | Yes | Yes | Yes |
| Were the outcome assessors blinded to the exposure status of participants? | CD | CD | CD | CD | No | CD | CD | CD |
| Was loss to follow-up after baseline 20% or less? | Yes | Yes | Yes | Yes | Yes | Yes | Yes | Yes |
| Were key potential confounding variables measured and adjusted statistically for their impact on the relationship between exposure(s) and outcome(s)? | Yes | Yes | Yes | Yes | Yes | Yes | Yes | Yes |
| **Quality Rating** | High | High | Good | High | Good | High | High | Good |
| **Risk of Bias** | Very low | Very low | Low | Very low | Low | Very low | Very low | Low |

CD, cannot determine; NA, not applicable; NR, not reported

| Criteria | Nakeshb<br>andi *et al.*<sup>75</sup> | Newton<br>*et al.*<sup>76</sup> | Olivas-Martínez *et al.*<sup>127</sup> | Ortiz-Brizuela<br>*et al.*<sup>77</sup> | Palaiodi<br>mos *et al.*<sup>78</sup> | Parker<br>*et al.*<sup>128</sup> | Parra-Bracamonte *et al.*<sup>129</sup> | Peng *et al.*<sup>138</sup> |
| --- | --- | --- | --- | --- | --- | --- | --- | --- |
| Was the research question or objective in this paper clearly stated? | Yes | Yes | Yes | Yes | Yes | Yes | Yes | Yes |
| Was the study population clearly specified and defined? | Yes | Yes | Yes | Yes | Yes | Yes | Yes | Yes |
| Was the participation rate of eligible persons at least 50%? | Yes | Yes | Yes | Yes | Yes | Yes | Yes | Yes |
| Were all the subjects selected or recruited from the same or similar populations (including the same time period)? Were inclusion and exclusion criteria for being in the study prespecified and applied uniformly to all participants? | Yes | Yes | Yes | Yes | Yes | Yes | Yes | Yes |
| Was a sample size justification, power description, or variance and effect estimates provided? | Yes | Yes | Yes | Yes | Yes | Yes | Yes | Yes |
| For the analyses in this paper, were the exposure(s) of interest measured prior to the outcome(s) being measured? | Yes | No | Yes | Yes | Yes | Yes | Yes | Yes |
| Was the timeframe sufficient so that one could reasonably expect to see an association between exposure and outcome if it existed? | No | No | No | No | No | No | No | No |
| For exposures that can vary in amount or level, did the study examine different levels of the exposure as related to the outcome (e.g., categories of exposure, or exposure measured as continuous variable)? | Yes | Yes | Yes | Yes | Yes | Yes | Yes | Yes |
| Were the exposure measures (independent variables) clearly defined, valid, reliable, and implemented consistently across all study participants? | Yes | Yes | Yes | No | Yes | Yes | Yes | Yes |
| Was the exposure(s) assessed more than once over time? | No | Yes | Yes | Yes | CD | CD | No | No |
| Were the outcome measures (dependent variables) clearly defined, valid, reliable, and implemented consistently across all study participants? | Yes | Yes | Yes | Yes | Yes | Yes | Yes | Yes |
| Were the outcome assessors blinded to the exposure status of participants? | CD | Yes | Yes | CD | Yes | CD | CD | CD |
| Was loss to follow-up after baseline 20% or less? | Yes | Yes | Yes | Yes | Yes | Yes | Yes | Yes |
| Were key potential confounding variables measured and adjusted statistically for their impact on the relationship between exposure(s) and outcome(s)? | Yes | Yes | Yes | CD | Yes | CD | Yes | Yes |
| **Quality Rating** | Good | High | High | Good | High | Good | Good | Good |
| **Risk of Bias** | Low | Very low | Very low | Low | Very low | Low | Low | Low |

CD, cannot determine; NA, not applicable; NR, not reported

| Criteria | Pepe *et al.*<sup>79</sup> | Petersen *et al.*<sup>80</sup> | Petrilli *et al.*<sup>81</sup> | Pettit *et al.*<sup>82</sup> | Philipos *e et al.*<sup>83</sup> | Pongpiru *l et al.*<sup>84</sup> | Rachel *et al.*<sup>103</sup> | Ramlall *et al.*<sup>85</sup> |
| --- | --- | --- | --- | --- | --- | --- | --- | --- |
| Was the research question or objective in this paper clearly stated? | Yes | Yes | Yes | Yes | Yes | Yes | Yes | Yes |
| Was the study population clearly specified and defined? | Yes | Yes | Yes | Yes | Yes | Yes | Yes | Yes |
| Was the participation rate of eligible persons at least 50%? | Yes | Yes | Yes | Yes | Yes | Yes | Yes | Yes |
| Were all the subjects selected or recruited from the same or similar populations (including the same time period)? Were inclusion and exclusion criteria for being in the study prespecified and applied uniformly to all participants? | Yes | Yes | Yes | Yes | Yes | Yes | Yes | Yes |
| Was a sample size justification, power description, or variance and effect estimates provided? | Yes | Yes | Yes | Yes | Yes | Yes | Yes | Yes |
| For the analyses in this paper, were the exposure(s) of interest measured prior to the outcome(s) being measured? | Yes | Yes | Yes | Yes | Yes | Yes | Yes | Yes |
| Was the timeframe sufficient so that one could reasonably expect to see an association between exposure and outcome if it existed? | No | No | No | No | No | Yes | CD | No |
| For exposures that can vary in amount or level, did the study examine different levels of the exposure as related to the outcome (e.g., categories of exposure, or exposure measured as continuous variable)? | Yes | Yes | Yes | Yes | Yes | Yes | Yes | Yes |
| Were the exposure measures (independent variables) clearly defined, valid, reliable, and implemented consistently across all study participants? | Yes | Yes | Yes | Yes | Yes | Yes | Yes | Yes |
| Was the exposure(s) assessed more than once over time? | No | Yes | No | No | No | Yes | Yes | NO |
| Were the outcome measures (dependent variables) clearly defined, valid, reliable, and implemented consistently across all study participants? | Yes | Yes | Yes | Yes | Yes | Yes | Yes | Yes |
| Were the outcome assessors blinded to the exposure status of participants? | CD | CD | NR | CD | CD | Yes | CD | CD |
| Was loss to follow-up after baseline 20% or less? | Yes | Yes | Yes | Yes | Yes | Yes | Yes | Yes |
| Were key potential confounding variables measured and adjusted statistically for their impact on the relationship between exposure(s) and outcome(s)? | Yes | Yes | Yes | CD | Yes | Yes | Yes | Yes |
| **Quality Rating** | Good | High | Good | Good | Good | High | High | Good |
| **Risk of Bias** | Low | Very low | Low | Low | Low | Very low | Very low | Low |

CD, cannot determine; NA, not applicable; NR, not reported

| Criteria | Rao *et al.*<sup>86</sup> | Reilev *et al.*<sup>87</sup> | Rodriguez *et al.*<sup>130</sup> | Rodríguez-Molinero *et al.*<sup>88</sup> | Salacup *G et al.*<sup>131</sup> | Rottoli *et al.*<sup>89</sup> | Shah *et al.*<sup>132</sup> | Shekhar *et al.*<sup>90</sup> | Simmonet *et al.*<sup>152</sup> |
| --- | --- | --- | --- | --- | --- | --- | --- | --- | --- |
| Was the research question or objective in this paper clearly stated? | Yes | Yes | Yes | Yes | Yes | Yes | Yes | Yes | Yes |
| Was the study population clearly specified and defined? | Yes | Yes | Yes | Yes | Yes | Yes | Yes | Yes | Yes |
| Was the participation rate of eligible persons at least 50%? | Yes | Yes | Yes | Yes | Yes | Yes | Yes | Yes | Yes |
| Were all the subjects selected or recruited from the same or similar populations (including the same time period)? Were inclusion and exclusion criteria for being in the study prespecified and applied uniformly to all participants? | Yes | Yes | Yes | Yes | Yes | Yes | Yes | Yes | Yes |
| Was a sample size justification, power description, or variance and effect estimates provided? | Yes | Yes | Yes | Yes | Yes | Yes | Yes | Yes | Yes |
| For the analyses in this paper, were the exposure(s) of interest measured prior to the outcome(s) being measured? | Yes | Yes | Yes | Yes | No | Yes | Yes | Yes | Yes |
| Was the timeframe sufficient so that one could reasonably expect to see an association between exposure and outcome if it existed? | CD | No | CD | No | Yes | No | No | No | No |
| For exposures that can vary in amount or level, did the study examine different levels of the exposure as related to the outcome (e.g., categories of exposure, or exposure measured as continuous variable)? | Yes | Yes | Yes | Yes | Yes | Yes | Yes | Yes | Yes |
| Were the exposure measures (independent variables) clearly defined, valid, reliable, and implemented consistently across all study participants? | Yes | Yes | Yes | Yes | Yes | No | Yes | Yes | Yes |
| Was the exposure(s) assessed more than once over time? | No | Yes | No | No | No | No | No | No | No |
| Were the outcome measures (dependent variables) clearly defined, valid, reliable, and implemented consistently across all study participants? | Yes | No | Yes | Yes | Yes | Yes | Yes | Yes | Yes |
| Were the outcome assessors blinded to the exposure status of participants? | Yes | CD | CD | CD | CD | CD | No | CD | Yes |
| Was loss to follow-up after baseline 20% or less? | Yes | Yes | Yes | Yes | Yes | Yes | Yes | Yes | Yes |
| Were key potential confounding variables measured and adjusted statistically for their impact on the relationship between exposure(s) and outcome(s)? | Yes | Yes | Yes | Yes | Yes | Yes | Yes | Yes | Yes |
| **Quality Rating** | High | Good | High | Good | Good | Good | High | Good | High |
| **Risk of Bias** | Very low | Low | Very low | Low | Low | Low | Very low | Low | Very low |

CD, cannot determine; NA, not applicable; NR, not reported

| Criteria | Steinberg *et al.*<sup>92</sup> | Suleyman *et al.*<sup>93</sup> | Tonetti *et al.*<sup>94</sup> | Vaquero-Roncer *et al.*<sup>96</sup> | Wang J *et al.*<sup>104</sup> | Wang Min *et al.*<sup>3</sup> | Wang R *et al.*<sup>97</sup> | Xiang *et al.*<sup>105</sup> | Zheng *et al.*<sup>98</sup> |
| --- | --- | --- | --- | --- | --- | --- | --- | --- | --- |
| Was the research question or objective in this paper clearly stated? | Yes | Yes | Yes | Yes | Yes | Yes | Yes | Yes | Yes |
| Was the study population clearly specified and defined? | Yes | Yes | Yes | Yes | Yes | Yes | Yes | Yes | Yes |
| Was the participation rate of eligible persons at least 50%? | Yes | Yes | Yes | Yes | Yes | Yes | Yes | Yes | Yes |
| Were all the subjects selected or recruited from the same or similar populations (including the same time period)? Were inclusion and exclusion criteria for being in the study prespecified and applied uniformly to all participants? | Yes | Yes | Yes | Yes | Yes | Yes | Yes | Yes | Yes |
| Was a sample size justification, power description, or variance and effect estimates provided? | Yes | Yes | Yes | Yes | Yes | Yes | Yes | Yes | Yes |
| For the analyses in this paper, were the exposure(s) of interest measured prior to the outcome(s) being measured? | Yes | No | Yes | Yes | Yes | Yes | Yes | Yes | Yes |
| Was the timeframe sufficient so that one could reasonably expect to see an association between exposure and outcome if it existed? | NO | No | No | No | No | NO | No | No | No |
| For exposures that can vary in amount or level, did the study examine different levels of the exposure as related to the outcome (e.g., categories of exposure, or exposure measured as continuous variable)? | Yes | Yes | Yes | Yes | Yes | Yes | yes | Yes | Yes |
| Were the exposure measures (independent variables) clearly defined, valid, reliable, and implemented consistently across all study participants? | Yes | Yes | Yes | Yes | Yes | Yes | Yes | Yes | Yes |
| Was the exposure(s) assessed more than once over time? | No | No | No | No | NO | No | No | No | No |
| Were the outcome measures (dependent variables) clearly defined, valid, reliable, and implemented consistently across all study participants? | Yes | Yes | Yes | Yes | No | Yes | Yes | Yes | Yes |
| Were the outcome assessors blinded to the exposure status of participants? | CD | CD | CD | Yes | CD | CD | CD | CD | CD |
| Was loss to follow-up after baseline 20% or less? | Yes | Yes | Yes | Yes | Yes | Yes | Yes | Yes | Yes |
| Were key potential confounding variables measured and adjusted statistically for their impact on the relationship between exposure(s) and outcome(s)? | No | Yes | Yes | Yes | Yes | CD | CD | CD | Yes |
| **Quality Rating** | Good | Good | Good | High | Good | Good | Good | Good | Good |
| **Risk of Bias** | Low | Low | Low | Very low | Low | Low | Low | Low | Low |

2B: Case control studies
| Criteria | Ciceri *et al.*<sup>112</sup> | Urra *et al.*<sup>95</sup> |
| --- | --- | --- |
| Was the research question or objective in this paper clearly stated and appropriate? | Yes | Yes |
| Was the study population clearly specified and defined? | Yes | Yes |
| Did the authors include a sample size justification? | Yes | Yes |
| Were controls selected or recruited from the same or similar population that gave rise to the cases (including the same timeframe)? | Yes | Yes |
| Were the definitions, inclusion and exclusion criteria, algorithms or processes used to identify or select cases and controls valid, reliable, and implemented consistently across all study participants? | Yes | Yes |
| Were the cases clearly defined and differentiated from controls? | Yes | Yes |
| If less than 100 percent of eligible cases and/or controls were selected for the study, were the cases and/or controls randomly selected from those eligible? | Yes | Yes |
| Was there use of concurrent controls? | Yes | No |
| Were the investigators able to confirm that the exposure/risk occurred prior to the development of the condition or event that defined a participant as a case? | Yes | Yes |
| Were the measures of exposure/risk clearly defined, valid, reliable, and implemented consistently (including the same time period) across all study participants? | Yes | Yes |
| Were the assessors of exposure/risk blinded to the case or control status of participants? | NR | NR |
| Were key potential confounding variables measured and adjusted statistically in the analyses? If matching was used, did the investigators account for matching during study analysis? | Yes | Yes |
| **Quality Rating** | High | High |
| **Risk of Bias** | Very low | Very low |

CD, cannot determine; NA, not applicable; NR, not reported

## Results

### Study characteristics of included studies

For the primary endpoint, severity, a total of 100 studies, consisting of 576,784 patients were included in the meta-analysis. The median age for included patients was 61.4 (55.3-65) with average 42.9% females (Table 1). Of the comorbidities considered, 29.4% were diabetics, 37.9% had heart diseases overall. Similarly, for the primary endpoint, i.e. disease severity, a total of seventy reports were included in the meta-analysis ^32, 42–110^. These had a combined sample size of 292,165 with 40,272 patients reaching the endpoint of high disease severity (Table 1). Similarly, a total of 51 studies were included for meta-analysis for the secondary outcome i.e. mortality^42, 45, 46, 48–52, 55, 63, 66, 75, 79, 80, 82, 85, 88, 89, 92, 101, 109, 111–140^. These had a combined sample size of 380,130 with 118,351 patients reaching the endpoint of mortality.

### Meta-analysis for severity outcome

Findings from the meta-analysis showed that being obese was correlated with increased severity of COVID 19 infections in comparison to non-obese patients (RR=1.46, 95% CI 1.34-1.60, p<0.001). Heterogeneity was high with I^2^ = 92 % (Figure 2).

**Figure 2:**
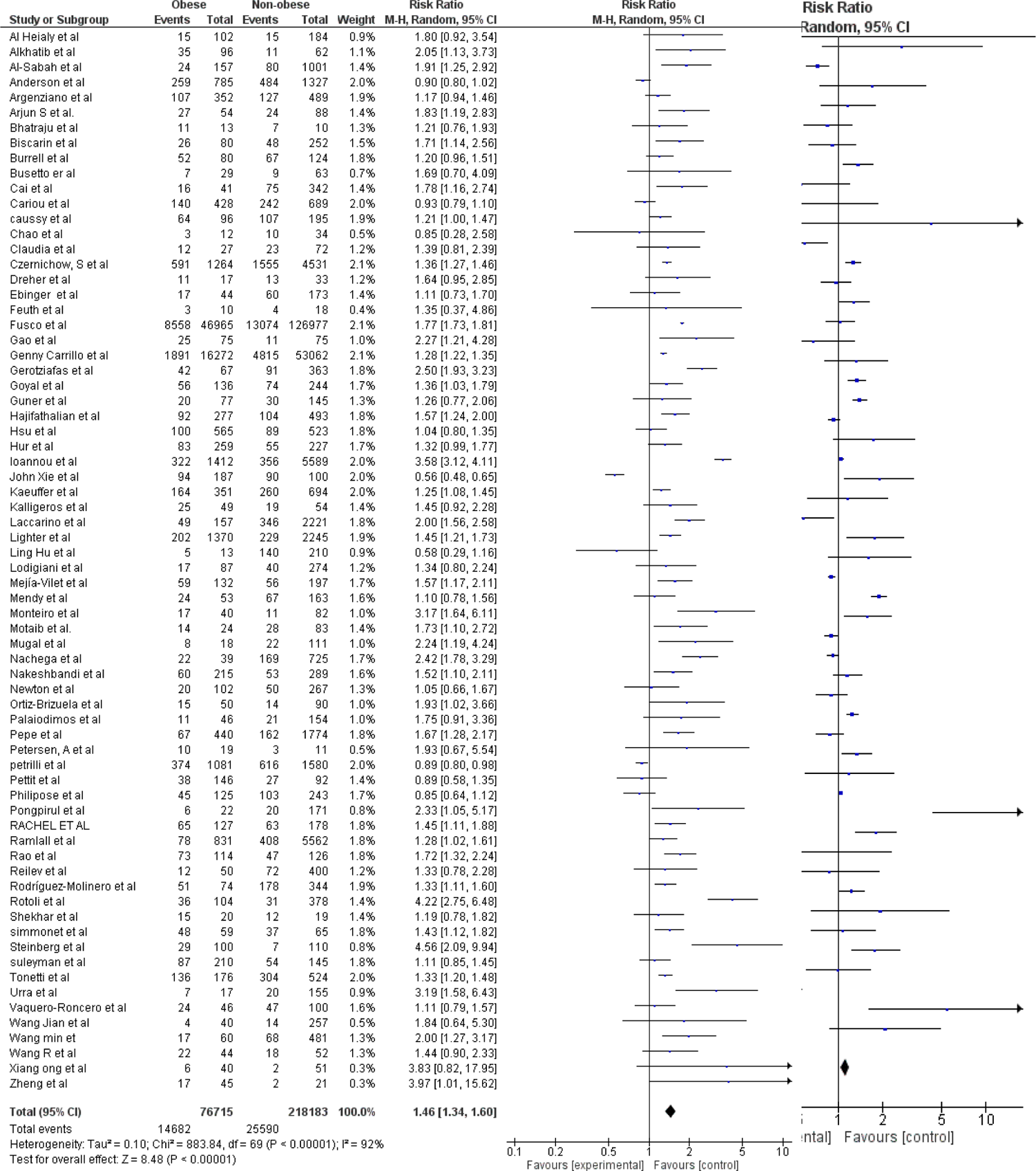
Forest plot for severity analysis

### Meta-analysis for mortality outcome

Meta-analysis findings showed that obesity was associated with increased risk of mortality from COVID 19 infections in comparison to non-obese patient population (RR=1.12, 95% CI 1.06-1.19, p<0.001). Heterogeneity was high with I^2^ = 88% (Figure 3).

**Figure 3:**
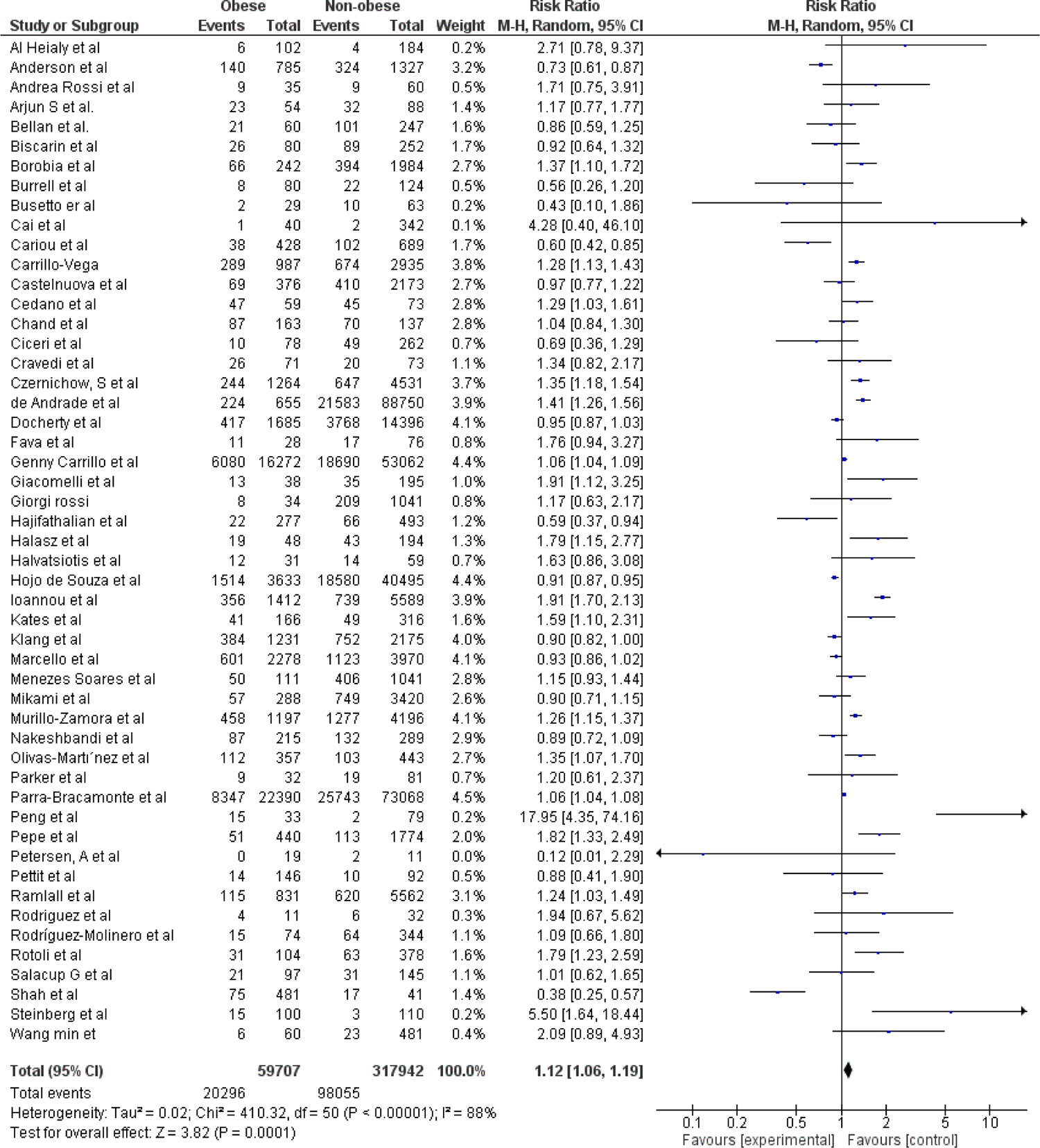
Forest plot for mortality analysis

### Multivariate meta-regression model for severity outcome

Multivariate meta-regression was performed to explain variations in the association between COVID-19 severity and obesity. We found female gender, pulmonary disease, diabetes, age, cardiovascular diseases, and hypertension covariates to be significant and this explained R^2^= 50% of the between-study heterogeneity in severity. The proportion of hypertension did not significantly affect the between-study variations and were therefore not included in the final equation. Figure 4 shows the resulting equation and individual covariate effect graphs.

**Figure 4:**
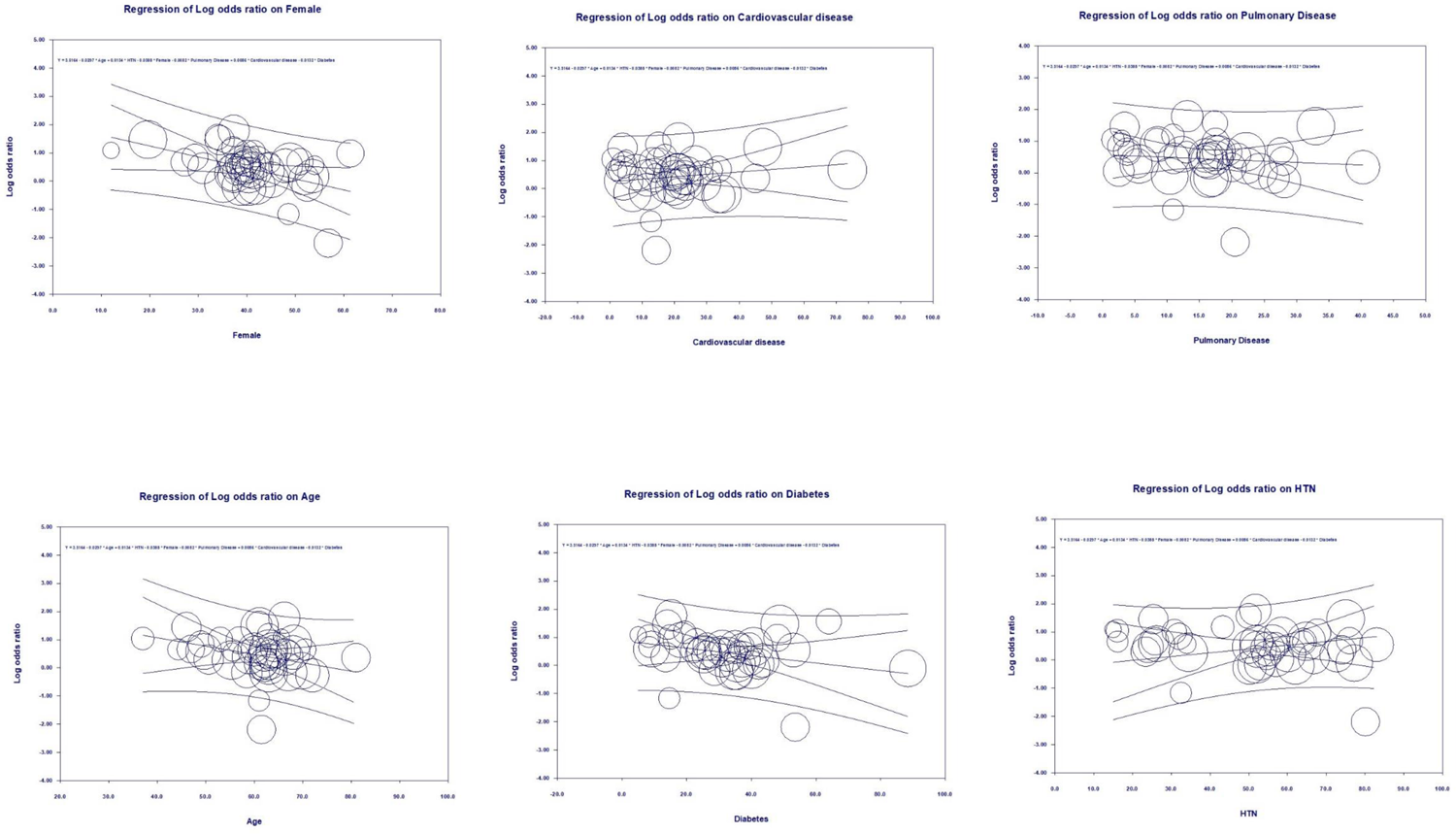
Severity meta-regression analysis

### Multivariate meta-regression model for mortality outcome

Multivariate meta-regression performed to explain variations in the association between mortality and obesity revealed that female gender, proportion of pulmonary disease, diabetes, hypertension, and cardiovascular diseases to be significant together. Overall, these covariates together explained R^2^=53% of the between-study heterogeneity in mortality. Figure 5 shows the resulting equation and individual covariate effect graphs.

**Figure 5:**
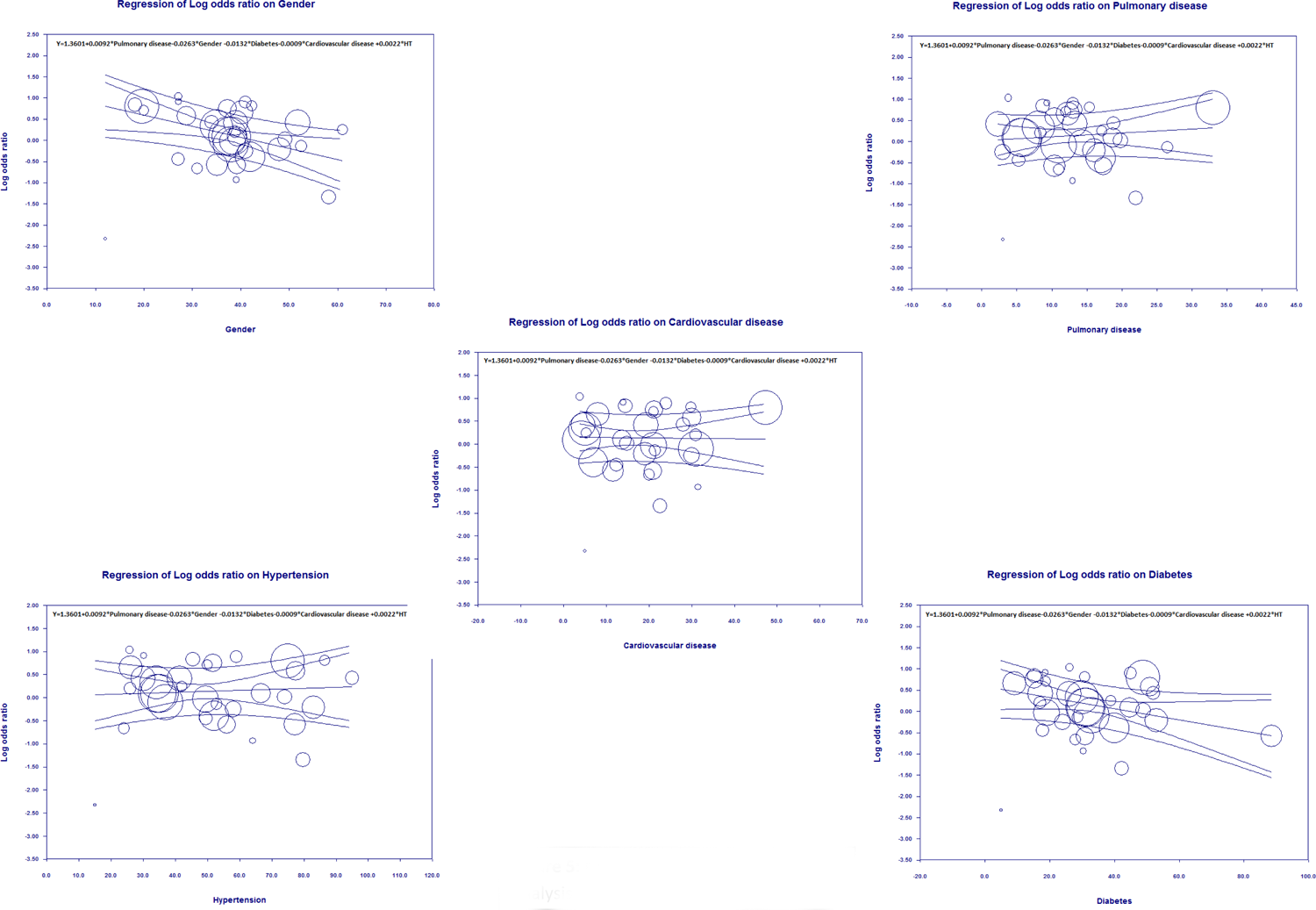
Mortality meta-regression analysis

### Publication Bias

Visual inspection of the standard error plots for the severity analysis also (Figure 7A) suggests symmetry without an underrepresentation of studies of any precision. However, in Egger’s regression test the null hypothesis of no small study effects was rejected at p<0.05 (estimated bias coefficient = −0.13 ± 0.42SE).

Similarly, visual inspection of the standard error plots for the mortality analysis (Figure 7B) suggests symmetry without an underrepresentation of studies of any precision. Corroborating inspection findings, Egger’s regression test, the null hypothesis of no small study effects, was rejected at p<0.05 (estimated bias coefficient = −0.17 ± 0.42SE).

### Sensitivity analysis

We did not find any statistical significance for risk of mortality as well as the risk for severity with COVID-19 when analyzed by BMI categories. However, we observed that underweight status (BMI<18 kg/m^2^) is associated with increased risk of mortality in COVID-19 (OR 1.52, 95% CI 1.19-1.94, p=<0.001; I^2^=0%) but not statistical significant to severity of COVID-19 (OR 1.10, 95% CI 0.81-1.48, p=0.54; I^2^=0%) as compared to normal BMI category of 18 kg/m^2^-25 kg/m^2^. We also did not observe any statistically significant changes while comparing BMI category 25 kg/m^2^ to 29.9 kg/m^2^ with respect to others in terms of mortality and severity of COVID-19 (Figure 6A-H).

**Figure 6:**
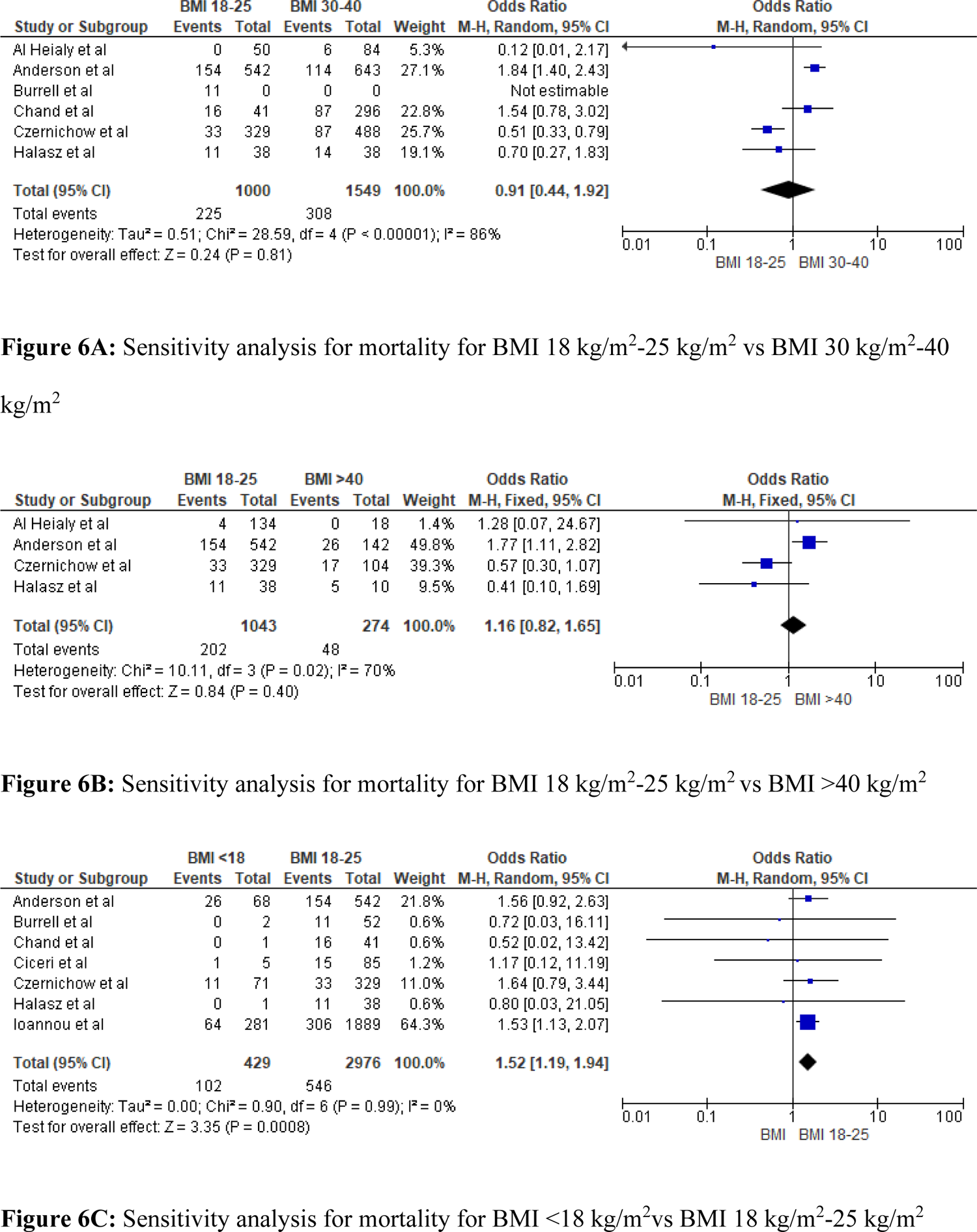
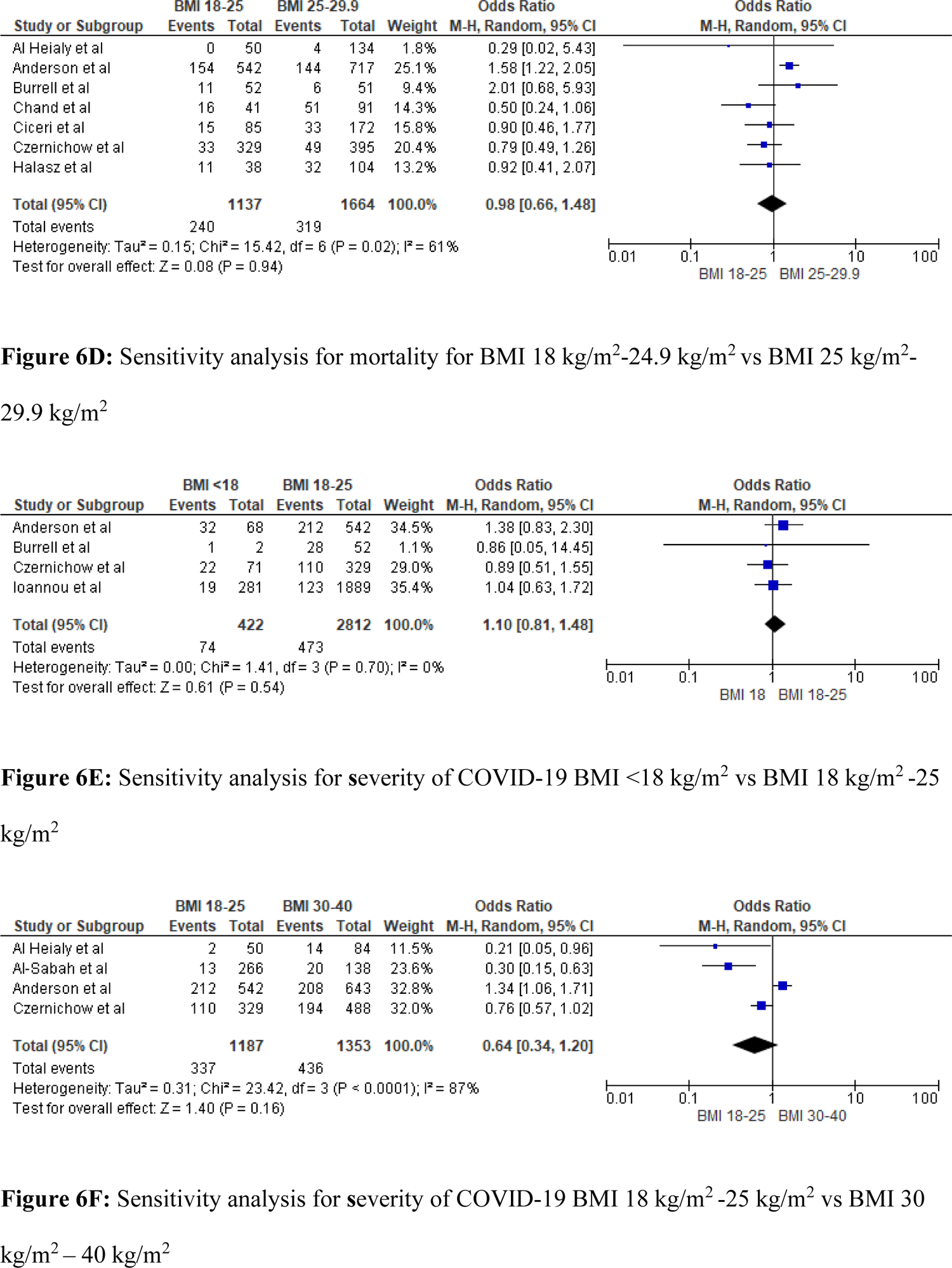
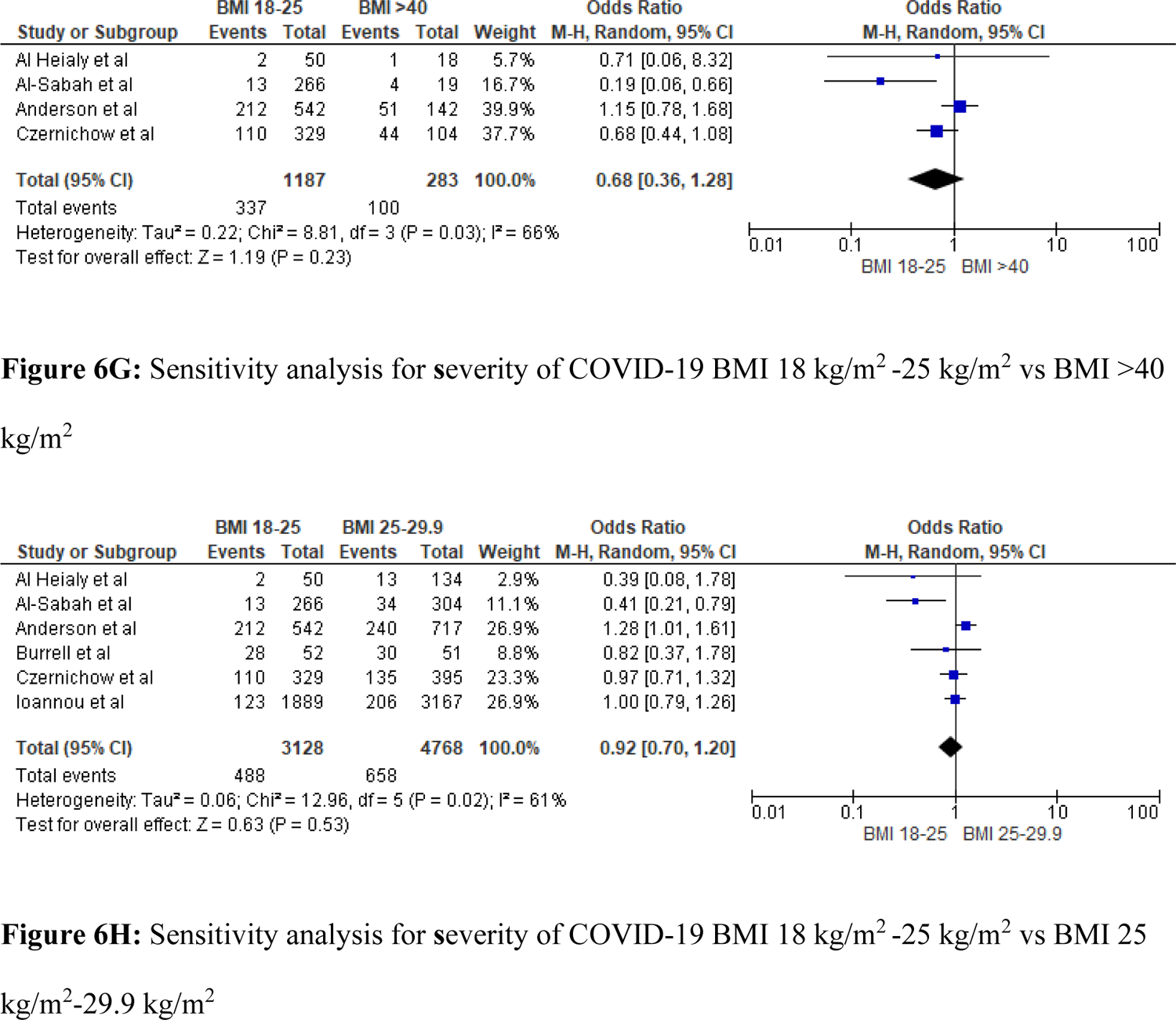
6A: Sensitivity analysis for mortality for BMI 18 kg/m^2^-25 kg/m^2^ vs BMI 30 kg/m^2^-40 kg/m^2^, **6B:** Sensitivity analysis for mortality for BMI 18 kg/m^2^-25 kg/m^2^ vs BMI >40 kg/m^2^, **6C:** Sensitivity analysis for mortality for BMI <18 kg/m^2^vs BMI 18 kg/m^2^-25 kg/m^2^, **6D:** Sensitivity analysis for mortality for BMI 18 kg/m^2^-24.9 kg/m^2^ vs BMI 25 kg/m^2^-29.9 kg/m^2^, **6E:** Sensitivity analysis for **s**everity of COVID-19 BMI <18 kg/m^2^ vs BMI 18 kg/m^2^ −25 kg/m^2^, **6F:** Sensitivity analysis for **s**everity of COVID-19 BMI 18 kg/m^2^ −25 kg/m^2^ vs BMI 30 kg/m^2^ – 40 kg/m^2^, **6G:** Sensitivity analysis for **s**everity of COVID-19 BMI 18 kg/m^2^ −25 kg/m^2^ vs BMI >40 kg/m^2^, **6H:** Sensitivity analysis for **s**everity of COVID-19 BMI 18 kg/m^2^ −25 kg/m^2^ vs BMI 25 kg/m^2^-29.9 kg/m^2^.

**Figure 7.**
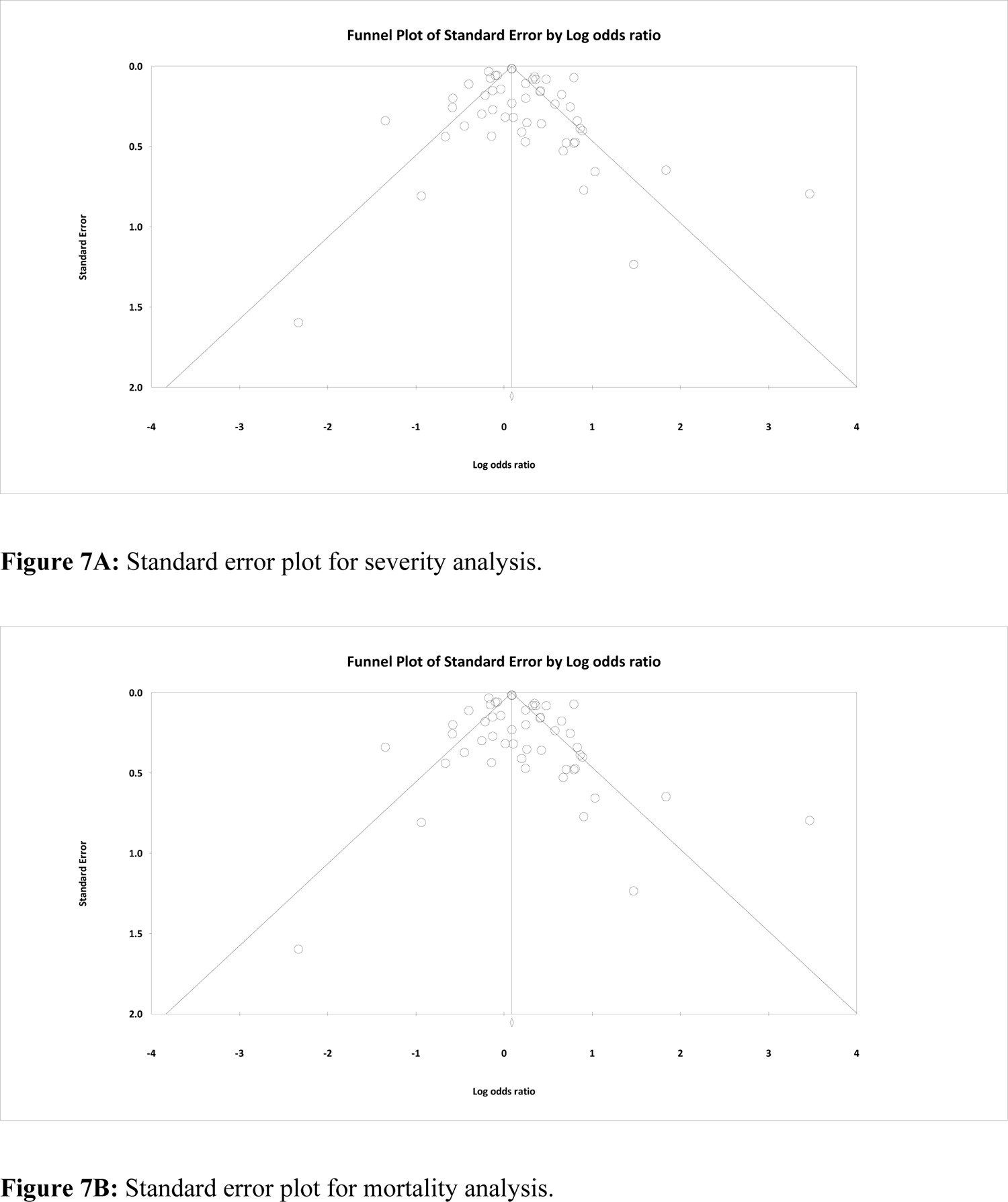
7A: Standard error plot for severity analysis, **7B:** Standard error plot for mortality analysis.

## Discussion

In this large meta-analysis with 100 studies, we found that obesity has a strong association with increased mortality & severity of COVID-19 infection. In addition, our meta-regression analysis suggests that obesity significantly increases the severity and mortality in COVID-19 patients.

Using a random effects model, we found that obese patients showed higher odds for mortality and severity i.e. ICU admissions or mechanical ventilation. Our results suggest that obese individuals are 1.5 times more likely to experience severe outcomes and 1.12 times more likely to die when compared to non-obese individuals with COVID-19 disease. Our meta-regression severity model suggested that 50% of the heterogeneity could be explained by age, gender, diabetes, hypertension, pulmonary and cardiovascular diseases. The mortality meta-regression model suggested that 53% of the heterogeneity could be accounted for by gender, diabetes, hypertension, pulmonary and cardiovascular diseases. Through these regression models, we were able to address major amount of heterogeneity seen in our meta-analysis.

In the existing literature, we found four meta-analysis (studies n=6, 17, 40, 76)^141–144^ that explored the association of obesity and worse outcomes in COVID-19 and found a similar association. On the contrary, one study refuted the possibility of this association. Owing to their small sample population (Studies n=2), it is likely that they were underpowered to tease out the true difference or association^145^. With a much larger sample size (n=100) our study provides a more robust evidence to establish this association.

Five meta-regression studies have evaluated the direct relationship between obesity and COVID-19 over the last year. Yang et al (studies n=41) concluded that, in COVID-19 patients, obesity is associated with increased mortality, increased rates of hospitalization, ICU admissions and the need for mechanical ventilation. However, they found no confounding factors causing heterogeneity in regards to hospitalization, ICU admission, and in-hospital mortality of COVID-19 patients^146^. In another such study, Mesas et al (studies n=60) described that obesity was linked to increased mortality only in studies with fewer chronic or critical patients and reported mean age of patients as the most important source of heterogeneity, followed by sex and health condition^147^. Soereto et al (studies n=16) reported that patients with higher BMI were at increased risk of developing ‘poor outcomes’ - defined as mortality, ICU admission, ARDS incidence, severe COVID-19, need for mechanical ventilation and hospitalization. In their meta regression, the heterogeneity in poor outcomes was explained by age, type 2 diabetes mellitus, hypertension, and gender^148^. Du et al and Chu et al (studies n=16 and 22) found that the association between obesity and COVID-19 severity and that with mortality was significantly influenced by age, but not with gender or other co morbidities^149, 150^. Our meta-regression identified the likely confounders to be age, gender, and co-morbidities such as diabetes, hypertension, pulmonary and cardiovascular diseases. Through this model, we were able to explain high heterogeneity with highest number of confounders, which other meta regression in the recent literature were not able to reach and define^146–150^. Thus, we were able to establish a strong association that obesity plays a remarkable role in worsening these outcomes in patients with COVID-19 infection. In the sensitivity analyses, we were only able to find statistically significant results for increased mortality in BMI<18 kg/m^2^ as compared to BMI 18 kg/m^2^-25 kg/m^2^, however, such significance was not noted in any other BMI categories with severity and mortality in COVID-19. This could be due to BMI being a very crude estimate of adiposity, may not be sensitive enough to tease out the real difference. Visceral adiposity would probably be a more reliable estimate to study these differences. However, in their study, Anderson et al. found that patients with obesity have a greater chance of intubation or mortality, with people with class 3 obesity having the greatest risk compared to overweight patients^45^.

Obesity is known to be associated with many adverse comorbid conditions^151^ including hypertension, atherogenic dyslipidemia, cardiovascular disease, insulin resistance or type 2 diabetes, altered cortisol metabolism, etc^152^. Obesity is associated with overexpression of ACE2 receptors and higher ACE2 receptors may aid infection and serve as viral reservoir ^153^ Moreover, obesity is known to be associated with endothelial dysfunction^154^, the key pathogenic event in COVID-19 infection leading to mortality and morbidity^155, 156^. Obesity or increased adiposity plays a key role in endothelial dysfunction by activating several cascade of pathological events namely-activation of renin-angiotensin system^157^, activation of procoagulant/hypercoagulation pathway ^158^, activation of proinflammatory mediators ^159^, insulin resistance ^160^, oxidative stress^161^, platelet dysfunction ^162^ and immune dysregulation ^163^. These events are summarized in Figure 8.

**Figure 8:**
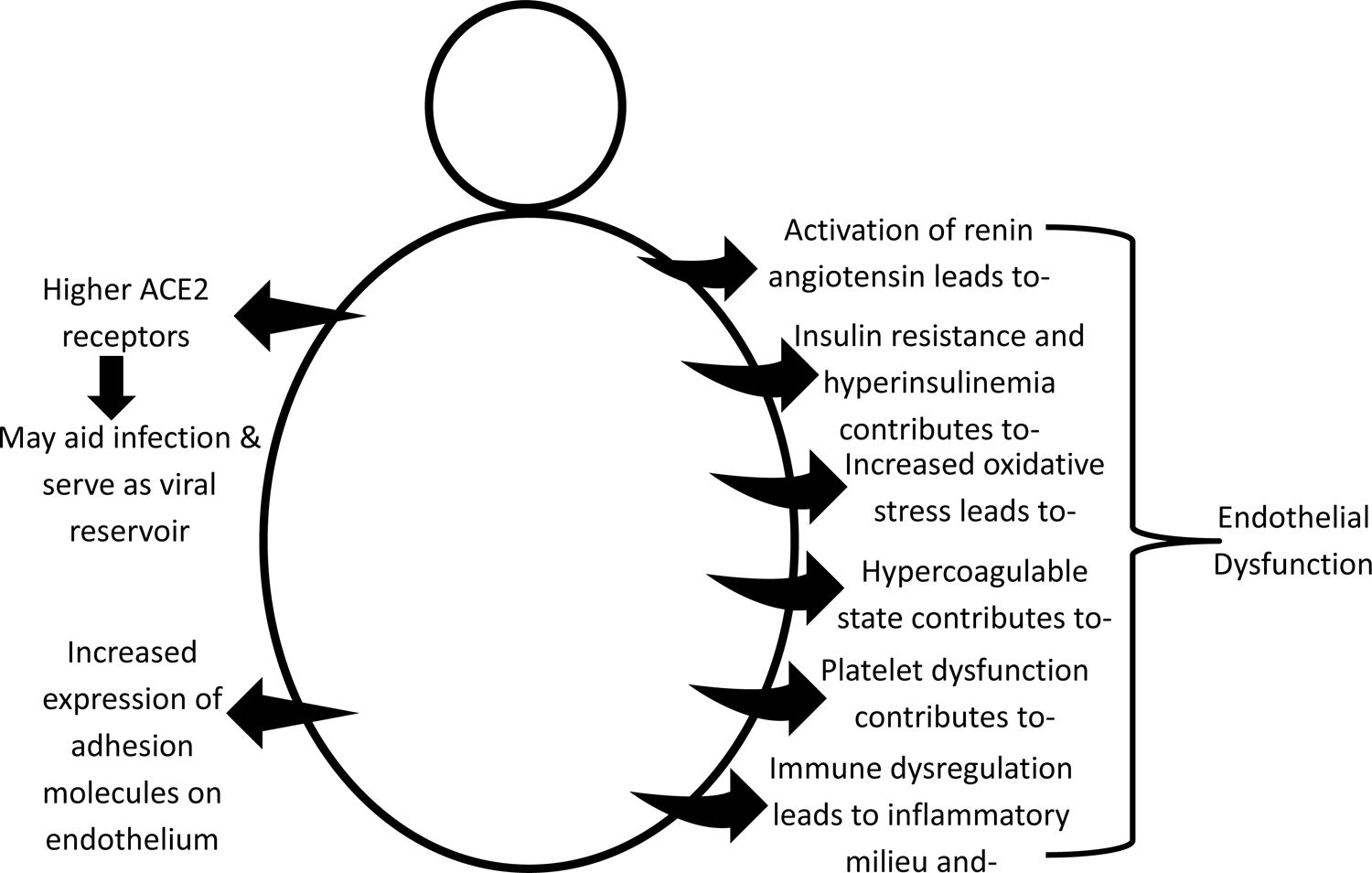
Several mechanisms of obesity’s role in endothelial dysfunction: A central event in pathogenicity of COVID-19 infection

In the study by Danzinger et al. obesity was found to be associated with increased incidence of acute kidney injury and increase in short- and long-term mortality^164^. Various meta-analyses were conducted to evaluate the association of obesity with mortality and severity of critically ill patients. The results were not universal, despite a wide variety of observations. In a total of 62,045 critically ill patients, Akinnusi et al compared the ICU mortality between obese and non-obese patients and found no dissimilarities^165^. Hogue et al. (n=22) conducted a meta-analysis of 88,051 patients and found that obesity did not impact ICU mortality^166^. However, Oliveros and Villamor et al. found that ICU mortality was increased only in underweight patients and reduced in overweight and obese patients^167^. In another study Zhao et al. observed that having a high BMI is related to longer duration on mechanical ventilation but lower mortality^168^. Therefore, it is unclear how obesity affects clinical outcomes in critically ill patients and more prospective studies are required to study the association between obesity and adverse outcomes in critical care.

The prime strength of this study is the large sample size. With an exhaustive search strategy, we compiled 100 studies conducted globally. We also added the most recent studies to our meta-analysis and meta-regression model including the studies that reported contradictory information. It enabled us to arrive at a more definitive conclusion about the risk associations. To define the heterogeneity in the meta-analysis, we also conducted a meta-regression analysis. For moderators, we used the most probable confounders based on the available evidence. This enabled us to delineate the impact of obesity as an independent risk factor for mortality and severity in COVID-19. However, our study is also subject to few limitations. We included five studies from preprint databases^71, 76, 83, 96, 101^ that may not be comparable to peer-reviewed articles in terms of their quality of methodology. However, in view of the time-sensitive nature of this pandemic, benefit of early dissemination of critical information and its inclusion in various analyses outweighs the risk from minor methodological flaws. Second factor was the heterogeneity in the studies in terms of the study design and methodology, patient sample and treatment received. There was a lack of uniformity in the type of outcomes evaluated for severity and their definitions in different studies. For the same reason, it was not possible to deduce the effect of obesity on the individual outcomes-ICU admission and mechanical ventilation. Third limitation is that the analysis was done with hospitalized patients only; hence we cannot generalize our results for patients seen in the outpatient clinic or treated at home. Analyzing outpatient data as well may help us to get the complete picture of the impact of obesity on the overall COVID-19 outcomes. Fourth limitation is that our analysis did not compare the outcomes with respect to visceral obesity and only BMI was used. However, it was beyond the scope of this analysis because of the lack of those details in most of the included studies. We suggest that prospective studies should obtain and report this information about their sample population.

Lastly, it is possible that some confounders which could have otherwise accounted for the residual heterogeneity were not evaluated in the meta-regression analysis due to limited information.

## Conclusion

In summary, our findings suggest that obesity significantly increases the risk of severity and mortality in hospitalized COVID-19 patients. Therefore, the inclusion of obesity or surrogate body mass index or visceral obesity in prognostic scores and streamlining the management strategy and treatment guidelines to account for the impact of obesity would be vital to improve patient outcomes in hospitalized COVID-19 patients. Our finding also serves as a call for the scientific community to further delve into its pathophysiology and identify potential pharmacological targets, since COVID-19 is an ever-evolving disease. Finally, this information must be disseminated to the general public to intensify the primary prevention of obesity.

## Data Availability

The data is available.

